# Automated, miniaturized, and scalable screening of healthcare workers, first responders, and students for SARS-CoV-2 in San Diego County

**DOI:** 10.1101/2021.06.25.21257885

**Authors:** Sydney C Morgan, Stefan Aigner, Catelyn Anderson, Pedro Belda-Ferre, Peter De Hoff, Clarisse A Marotz, Shashank Sathe, Mark Zeller, Noorsher Ahmed, Xaver Audhya, Nathan A Baer, Tom Barber, Bethany Barrick, Lakshmi Batachari, Maryann Betty, Steven M Blue, Brent Brainard, Tyler Buckley, Jamie Case, Anelizze Castro-Martinez, Marisol Chacón, Willi Cheung, LaVonnye Chong, Nicole G Coufal, Evelyn S Crescini, Scott DeGrand, David P Dimmock, J Joelle Donofrio-Odmann, Emily R Eisner, Mehrbod Estaki, Lizbeth Franco Vargas, Michele Freddock, Robert M Gallant, Andrea Galmozzi, Nina J Gao, Sheldon Gilmer, Edyta M Grzelak, Abbas Hakim, Jonathan Hart, Charlotte Hobbs, Greg Humphrey, Nadja Ilkenhans, Marni Jacobs, Christopher A Kahn, Bhavika K Kapadia, Matthew Kim, Sunil Kurian, Alma L Lastrella, Elijah S Lawrence, Kari Lee, Qishan Liang, Hanna Liliom, Valentina Lo Sardo, Robert Logan, Michal Machnicki, Celestine G Magallanes, Clarence K Mah, Denise Malacki, Ryan J Marina, Christopher Marsh, Natasha K Martin, Nathaniel L Matteson, Daniel J Maunder, Kyle McBride, Bryan McDonald, Daniel McDonald, Michelle McGraw, Audra R Meadows, Michelle Meyer, Amber L Morey, Jasmine R Mueller, Toan T Ngo, Julie Nguyen, Viet Nguyen, Laura J Nicholson, Alhakam Nouri, Victoria Nudell, Eugenio Nunez, Kyle O’Neill, R Tyler Ostrander, Priyadarshini Pantham, Samuel S Park, David Picone, Ashley Plascencia, Isaraphorn Pratumchai, Michael Quigley, Michelle Franc Ragsac, Andrew C Richardson, Refugio Robles-Sikisaka, Christopher A Ruiz, Justin Ryan, Lisa Sacco, Sharada Saraf, Phoebe Seaver, Leigh Sewall, Elizabeth W Smoot, Kathleen M Sweeney, Chandana Tekkatte, Rebecca Tsai, Holly Valentine, Shawn Walsh, August Williams, Min Yi Wu, Bing Xia, Brian Yee, Jason Z Zhang, Kristian G Andersen, Lauge Farnaes, Rob Knight, Gene W Yeo, Louise C Laurent

**Author notes:** These authors contributed equally to this work (Sydney C Morgan, Stefan Aigner, Catelyn Anderson, Pedro Belda-Ferre, Peter De Hoff, Clarisse A Marotz, Shashank Sathe, Mark Zeller). Senior authors (Kristian G Andersen, Lauge Farnaes, Rob Knight, Gene W Yeo, Louise C Laurent).

## Abstract

**Background:** Successful containment strategies for SARS-CoV-2, the causative virus of the COVID-19 pandemic, have involved widespread population testing that identifies infections early and enables rapid contact tracing. In this study, we developed a rapid and inexpensive RT-qPCR testing pipeline for population-level SARS-CoV-2 detection, and used this pipeline to establish a clinical laboratory dedicated to COVID-19 testing at the University of California San Diego (UCSD) with a processing capacity of 6,000 samples per day and next-day result turnaround times.

**Methods and findings:** Using this pipeline, we screened 6,786 healthcare workers and first responders, and 21,220 students, faculty, and staff from UCSD. Additionally, we screened 6,031 preschool-grade 12 students and staff from public and private schools across San Diego County that remained fully or partially open for in-person teaching during the pandemic. Between April 17, 2020 and February 5, 2021, participants provided 161,582 nasal swabs that were tested for the presence of SARS-CoV-2. Overall, 752 positive tests were obtained, yielding a test positivity rate of 0.47%. While the presence of symptoms was significantly correlated with higher viral load, most of the COVID-19 positive participants who participated in symptom surveys were asymptomatic at the time of testing. The positivity rate among preschool-grade 12 schools that remained open for in-person teaching was similar to the positivity rate at UCSD and lower than that of San Diego County, with the children in private schools being less likely to test positive than the adults at these schools.

**Conclusions:** Most schools across the United States have been closed for in-person learning for much of the 2020-2021 school year, and their safe reopening is a national priority. However, as there are no vaccines against SARS-CoV-2 currently available to the majority of school-aged children, the traditional strategies of mandatory masking, physical distancing, and repeated viral testing of students and staff remain key components of risk mitigation in these settings. The data presented here suggest that the safety measures and repeated testing actions taken by participating healthcare and educational facilities were effective in preventing outbreaks, and that a similar combination of risk-mitigation strategies and repeated testing may be successfully adopted by other healthcare and educational systems.

## Introduction

<u>Co</u>rona<u>vi</u>rus <u>D</u>isease 20<u>19</u> (COVID-19) is a serious respiratory illness caused by the <u>S</u>evere <u>A</u>cute <u>R</u>espiratory <u>S</u>yndrome <u>Co</u>rona<u>v</u>irus <u>2</u> (SARS-CoV-2). The COVID-19 pandemic has had a dramatic and negative impact on the health, well-being, and productivity of communities around the world, with over 165 million confirmed cases and 3.4 million deaths worldwide, including more than 33 million confirmed cases and 587,000 deaths in the United States (US) alone, within the 18 months since this virus was discovered (1).

Given the lack of widely available and highly effective preventative or therapeutic agents besides the recent ramp-up of vaccine availability, social interventions such as “stay-at-home” orders, temporary school and business closures, and mask-wearing/physical distancing measures have been enacted in much of the world to slow the spread of the SARS-CoV-2 virus. While many of these measures are effective at slowing viral spread through a community when carried out correctly (2–4), they do not completely halt the spread of the virus, nor are they sustainable long term. Indeed, a major concern in the US and other similarly affected countries is how to safely reopen schools. Some schools and universities in the US have partially reopened, using a hybrid of online and in-person teaching, or have fully reopened with increased safety measures in place. The CDC has recently issued recommendations on how to limit the spread of COVID-19 in schools (https://www.cdc.gov/coronavirus/2019-ncov/community/schools-childcare/operation-strategy.html), which include universal and correct use of masks, physical distancing, handwashing, and contact tracing, but, so far, few studies have investigated how these measures may work in practice in this environment (5–8).

The countries that have demonstrated the greatest success in controlling the COVID-19 pandemic, including New Zealand, Australia, and South Korea, have combined risk-mitigating strategies with large-scale and widespread testing, as well as aggressive contact tracing, to both identify outbreaks and curb community transmission. Importantly, this broad testing must be implemented with a minimal turnaround time to allow for effective contact tracing and isolation of affected persons. Even now, nearly 1.5 years after the initial reports of COVID-19 infections, it can be difficult for many people in the US and other countries to obtain a test for SARS-CoV-2 if they do not exhibit symptoms or have not been in known contact with a person with a confirmed case of COVID-19. In the context of overall limited testing capacity, these measures are intended to allow medical doctors to triage patients into appropriate treatment pipelines. However, in the context of a pandemic, these measures exclude key demographics, namely asymptomatic and pre-symptomatic carriers, who contribute to the “silent spread” of the virus (9, 10). Therefore, the current symptom- and contact-based testing strategies employed in much of the world are both unlikely to accurately capture the full extent to which this novel coronavirus can spread throughout our communities.

Some regions have screened large, representative proportions of their target populations with great success, and provide an insight into the proportion of COVID-19 cases that may be missed with current testing restrictions. These studies, which estimate that the rate of asymptomatic COVID-19 infection may be as high as 40-45%, highlight the importance of routine asymptomatic testing (11–13). Additionally, we now know that asymptomatic and pre-symptomatic infections are transmissible (14, 15).

The objective of this study was to develop and establish an accurate, high-throughput, rapid, and inexpensive SARS-CoV-2 screening pipeline for use at the population level. To this end, we performed SARS-CoV-2 screening on over 5,000 healthcare workers from two large healthcare systems in San Diego County and 1,162 first responders from San Diego Fire and Rescue. Additionally, we screened 21,220 students, faculty, and staff from the University of California, San Diego (UCSD), and 6,031 students and staff from preschool-grade 12 schools across San Diego County that had remained fully or partially open for in-person teaching throughout the COVID-19 pandemic (4,750 from 11 private schools and 1,281 from 13 public schools). We developed a RT-qPCR testing pipeline that included the miniaturization of a testing kit that was granted Emergency Use Authorization by the U.S. Food and Drug Administration. This pipeline was then used to establish a Clinical Laboratory Improvement Amendments (CLIA) certified laboratory at UCSD, and further refinements were made to increase throughput capacity to 6,000 samples per day with next-day results, and to enable accurate detection from self-collected anterior-nares swab samples. A secondary objective of this study was to identify asymptomatic and pre-symptomatic infections, and to evaluate the effectiveness of health and safety measures implemented by healthcare and educational facilities in response to the COVID-19 pandemic, particularly those developed at educational facilities to safely re-open schools for in-person learning.

## Methods

### SEARCH Study

The SEARCH study (<u>S</u>an Diego <u>E</u>pidemiology <u>a</u>nd <u>R</u>esearch Study for <u>C</u>OVID-19 <u>H</u>ealth) was a multi-site study whose aim was to evaluate the presence of COVID-19 infection in potentially exposed healthcare worker and first responder populations in San Diego County (SD County). In particular, employees from Rady Children’s Hospital San Diego (RCHSD), Rady Children’s Institute for Genomic Medicine (RCIGM), Scripps Health, and the San Diego Fire- Rescue Department (SDFD) were screened, along with a small number of employees from other institutions who heard of the study by word of mouth and were allowed to participate. These other institutions included the University of California, San Diego (UCSD), Children’s Primary Care Medical Group, Rady Children’s Specialists of San Diego, and Sharp HealthCare. Screening took place from April 17 through June 30, 2020. Nasopharyngeal (NP) swabs were obtained from study participants by trained healthcare workers, and RT-qPCR testing was performed in collaborating basic science labs at UCSD and Scripps Research to test for the presence of SARS-CoV-2 in the samples. Participant samples that tested positive using the research lab screening protocol were subjected to confirmatory clinical tests, at which point the positive results were reported to both the survey participant and the SD County Department of Public Health. The members of the research team had final responsibility for the survey design, clinical protocol, and trial oversight. The UCSD Institutional Review Boards (IRB) provided human subject protection oversight of the SEARCH study (IRB approval #200470).

### Nasopharyngeal swab production

Due to a shortage of NP swabs when the SEARCH study began, 3D-printed swabs were designed and printed for testing. The swab shafts were 3D-printed in nylon, utilizing material extrusion (i.e. fused filament fabrication) on an Onyx One Professional Desktop 3D Printer (Markforged, Watertown, MA, USA). The nylon swab tips were wrapped with rayon fibers and steam-sterilized. While rayon fibers have previously been found to be slightly inferior to flocked nylon (16), they were readily available and more easily manufactured in this setting.

We validated the performance of these 3D-printed swabs in parallel with commercially available NP swabs (FLOQSwab, Copan Diagnostics Inc., Murrieta, CA, USA), using both a commercially available standard viral respiratory panel in pediatric patients, as well as SARS-CoV-2 testing in known positive adult patients. The detailed protocol and complete validation information has been made available on protocols.io (17). Briefly, at Rady Children’s Hospital San Diego (RCHSD, CA, USA), 25 pediatric patients were swabbed by respiratory therapists using both the 3D-printed swabs and the commercially-sourced swabs, of which 22 samples were run on the ePlex Respiratory Pathogen (RP) Panel (GenMark Dx, Carlsbad, CA, USA), which test for viruses including adenovirus, human metapneumovirus, respiratory syncytial virus (RSV), and variants of influenza and parainfluenza. The remaining three samples, two negative and one positive for SARS-CoV-2, were analyzed using the Simplexa COVID-19 Direct Kit (DiaSorin Molecular, Cypress, CA, USA). Finally, nine adult outpatients who had previously tested positive for SARS-CoV-2 using various FDA-approved tests performed self-swabs 3 to 14 days later, some patients with paired parallel commercial and 3D printed swabs (*n* = 4) and others with the 3D-printed swabs alone (*n* = 5). Results from the 3D-printed swab validation are presented in Table S1.

### Sample collection and population screening

Inclusion criteria for this study included: (1) Individuals 18 years of age or older with possible exposure to COVID-19 in a work setting. Exclusion criteria included: (1) individuals from whom it was not possible to obtain an adequate nasopharyngeal swab; (2) individuals who needed immediate medical intervention.

In an effort to include healthcare workers throughout SD County, multiple study sites at hospitals and clinics were set up; each study site was staffed by individuals from that hosting location. A fixed testing location was set up at RCHSD and a mobile testing unit was created to serve nine Scripps Health locations (Scripps Green Hospital; Scripps Memorial La Jolla Hospital; Scripps Memorial Encinitas Hospital; Scripps Mercy San Diego Hospital; Scripps Mercy Chula Vista Hospital; Scripps Clinic Rancho Bernardo; Scripps Coastal Vista Center; Scripps Clinic Mission Valley; and Scripps Campus Point).

Potential healthcare worker participants were informed of the study through an organizational all-user email communication as well as a flyer that was sent to Rady and Scripps Health employees and which contained a QR code that allowed them to fill out an electronic consent form and symptom questionnaire prior to arrival at a study site using REDCap (Research Electronic Data Capture) software hosted at RCHSD. REDCap is a secure, web-based software platform designed to support data capture for research studies (18, 19).

Paper versions of these forms were also available at the study site if an eligible participant was not able to complete them electronically. Potential firefighter and lifeguard participants were informed of the study through an assignment that was posted to every SDFD worker on their Target Solutions (Vector Solutions, Tampa, FL, USA) accounts. Target Solutions is an online training management system which requires assignments to be opened, read, and confirmed prior to the assignment deadline, to ensure all personnel receive and acknowledge their contents. The assignment contained the details of the study as well as instructions on how to participate. In this way, all firefighters and lifeguards in the SDFD were informed of this study.

Potential participants were then asked to complete a three-question quiz: (1) Is participation in this study voluntary? (Answer: yes); (2) Will participants be told about their study findings? (Answer: not necessarily); (3) Is participating in this study the same as receiving a clinical test? (Answer: no). Those who answered any questions incorrectly were re-educated by study staff prior to final enrollment and the collection of samples. The Enrollment Screening Form - completed either electronically using a secure REDCap survey, or using a paper version - collected identifiable information including name, address, contact phone number, date of birth, workplace information, and date and site of testing, in addition to information regarding comorbidities and any active symptoms on the day of testing. A barcode was entered into an electronic data capture file for each participant with matching labels placed on a participant information sheet as well as on the NP swab collection tube, in order to link each sample with the participant. A barcode-associated “synthetic name” was also generated, and was cross-checked with each participant at registration and again at the swabbing station to prevent sample mix-up. Protected health information (PHI) was not provided to the research testing laboratories. Instead, names and identifiable information were stored on REDCap at RCHSD, as specified in the IRB protocol (IRB approval #200470).

An NP swab sample was collected from each participant by sampling the posterior nasopharynx through both nares, according to instructions for NP swab collection from the CDC (https://www.cdc.gov/flu/pdf/professionals/flu-specimen-collection-poster.pdf). A qualified team member with appropriate personal protective equipment (PPE), consisting of gown, gloves, N95 respirator, and face shield, obtained the sample and placed the NP swab into the barcode-labeled collection tube containing 3 mL viral transport medium (VTM), which was prepared according to CDC guidelines (https://www.cdc.gov/coronavirus/2019-ncov/downloads/Viral-Transport-Medium.pdf). At the end of each sampling day, samples were transported on wet ice in a cooler to a Biosafety Laboratory 2 Plus (BSL-2+) laboratory at Scripps Research (La Jolla, CA, USA) for sample accessioning and plating, as described in the Sample processing, RNA extraction, and RT-qPCR section below.

At RCHSD, NP sample collection of healthcare workers and firefighters for this study involved 192 screening hours over 33 sampling days, between April 17 and June 30, 2020, with two registered nurses (RNs) involved in consenting participants for the study, two respiratory therapists involved in swabbing participants, and two additional RNs acting as support staff involved in participant information data entry and specimen handling support, for a total of six healthcare workers at each screening site on each day. Across all the Scripps Health locations, NP sample collection for this study involved 76 screening hours over 20 sampling days, between April 24 and June 30, 2020. The mobile sample collection team increased from six to ten staff (four additional support staff for data entry and specimen handling) for locations with high demand. For the SDFD employees, NP samples were collected at the permanent sampling location at RCHSD between June 1 and June 30, 2020.

### INSPECT application development

Specimens and results were tracked through the processing pipeline using the INSPECT (<u>In</u>stant <u>S</u>ervice <u>P</u>latform for <u>E</u>mergency <u>C</u>OVID-19 <u>T</u>ests) application. This Laboratory Information Management System (LIMS) was designed specifically for tracking SARS-CoV-2 tests, and includes tracking of specimen ID, plate layouts, plate barcodes, RT-qPCR test results, and participant IDs. All code and protocols needed to create a lab-tailored version of the INSPECT app are freely available on GitHub (https://github.com/SEARCH-Alliance/inspect) and protocols.io (20).

### SEARCH sample processing, RNA extraction, and RT-qPCR

Sample processing and viral RNA extraction was performed using a KingFisher^TM^ Flex automated sample preparation machine (Thermo Fisher, Waltham, MA, USA) and an Eppendorf epMotion® 5070 automated liquid handling workstation (Enfield, CT, USA). RNA was extracted with the Omega Bio-tek Mag-Bind Viral DNA/RNA kit. A detailed protocol of the RNA extraction process used in the SEARCH study is available on protocols.io (21). Following RNA extraction of samples, SARS-CoV-2 detection was performed using a miniaturized and automated RT- qPCR procedure. Viral RNA was detected using the TaqPath COVID-19 Combo Kit (ThermoFisher Scientific, Waltham, MA, USA). This viral RNA detection kit is approved by the Emergency Use Authorization (EUA) authority of the US FDA for the detection of SARS-CoV-2, and targets three regions of the SARS-CoV-2 genome - N gene, S gene, and ORF1ab gene - as well as an internal positive control, MS2. The miniaturization of the RT-qPCR process (from the standard 25 µL to a 3 µL reaction volume) involved the use of two Mosquito robotic liquid handlers (STP Labtech Ltd., formerly TTP Labtech Ltd., Boston, MA, USA): a 16-channel liquid handler (HV Genomics), and an HV X1 hit/cherry picker single-channel liquid handler. RT-qPCR was performed on a QuantStudio™ 5 Real-Time PCR System (Applied Biosystems). A detailed protocol of the miniaturized and automated RT-qPCR process used in this study is available on protocols.io (22). Using the TaqPath COVID-19 Multiplex Real-Time RT-PCR Assay, samples are considered positive when at least two out of three viral target genes (S gene, N gene, and ORF1ab) amplify using a threshold cycle (Ct) of 37 cycles, and samples are considered negative when none of the three viral target genes amplify but the internal control (MS2) does amplify. Inconclusive test results are those in which only one of three viral target genes amplifies, while invalid tests are those in which no viral target genes amplify, and the internal control also fails to amplify. Samples that produced inconclusive and invalid results from the initial analysis were re-extracted and re-amplified, and the decision tree recommended by the manufacturer (Thermo Fisher) was used to determine the final result; data presented here are the final results obtained after re-extraction and re-amplification, in cases where this was necessary.

### Validation of RT-qPCR miniaturization and research workflow

A technical validation of the RT-qPCR miniaturization was performed by comparing the miniaturization reactions to full-scale reactions. RT-qPCR performance of the miniaturized reactions was compared to published results for full-scale reactions to evaluate equivalency (Fig S1). The limit of detection (LOD) was calculated for the miniaturized reactions using two different methods: (1) SARS-CoV-2 viral RNA was spiked into the RT-qPCR reaction in different concentrations (1-128 viral genome equivalents per reaction) along with RNA extracted from negative NP control samples; (2) SARS-CoV-2 viral RNA was spiked into negative NP control samples in lysis buffer, before RNA extraction, in different concentrations (1-100,000 viral genome equivalents/mL input sample) (Fig S2). RT-qPCR performance of the miniaturized reactions were found to be equivalent to the full-scale reactions (Fig S1). The limit of detection for the miniaturized reactions (using the evaluation criteria recommended by the manufacturer, and requiring all three replicates to show a positive result) was found to be 500 viral RNA copies/mL input sample, with >4 viral RNA copies per RT-qPCR reaction (Fig S2).

Samples that tested positive through the research pipeline were subjected to confirmatory clinical tests by a collaborating Clinical Laboratory Improved Amendments (CLIA) lab at UCSD. A number of negative samples were also sent to the collaborating CLIA lab to confirm their negative status as determined through the research pipeline. In total, 21 positive and 141 negative samples obtained through the research pipeline were re-tested by the collaborating CLIA lab: all 141 negative samples were confirmed negative, and 18/21 positive samples were confirmed as true positives, with 3/21 samples returning a false-positive result. The true positives were reported to the study participants and the SD County Public Health Department.

### EXCITE Lab

After the SEARCH study (the initial proof-of-concept study) was complete, the methodology developed by the SEARCH team was adapted to establish the <u>Ex</u>pedited <u>C</u>OVID-19 Identification <u>E</u>nvironment (EXCITE) lab, a CLIA-certified laboratory at UCSD dedicated to COVID-19 testing. The EXCITE lab was developed to provide rapid, asymptomatic population screening for UCSD students/faculty/staff, and also partnered with local public and private preschool-grade 12 schools and SDFD to test their populations. The UCSD Institutional Review Boards (IRB) provided human subject protection oversight of the data obtained by the EXCITE lab (IRB approval #210817X).

### Translation of the research workflow into the CLIA environment

The research workflow from the SEARCH study was adapted for clinical use by the EXCITE lab, with the goal of repeated screening of large populations as a component of programs developed to safely increase educational and business activities during the COVID-19 pandemic. Five major changes to the SEARCH pipeline were implemented by the EXCITE lab to improve patient acceptability, enhance the safety of testing personnel, increase assay throughput, ensure compliance with College of American Pathologists (CAP) and CLIA regulations, and enable reporting of clinical results to patients’ medical records and to the required public health agencies.

First, we validated anterior nares (AN) sample collection via self-swabbing. NP swabs are considered the gold standard sample type used to test for SARS-CoV-2. However, these swabs are uncomfortable for the patient and require collection by a clinical provider at close proximity, which decreases compliance and increases cost and risk, making it infeasible for large-scale repeated screening programs. Anterior nares swabs can be self-collected with minimal discomfort, while providing comparable test results.

Second, to enable safer handling of large numbers of samples, we changed from sample collection in VTM, in which viral particles remain infectious, to PrimeStore® Molecular Transport Medium (MTM), which immediately inactivates the SARS-CoV-2 virus. Following validation with MTM, we additionally validated sample collection in Mawi DNA Technologies iSWAB Microbiome buffer (Mawi) as well as 5% sodium dodecyl sulfate (SDS). AN swabs in MTM, Mawi, and SDS are referred to here as “ANM”, “ANW”, and “AND”, respectively. All three media types inactivate the virus, allowing for self-collection and safer handling, but Mawi and SDS have the additional benefit of being non-toxic substances, allowing for unsupervised self- collectio n of samples.

Third, to increase assay throughput, we centralized our operations from three research laboratories located ∼10 min drive apart into two adjacent rooms in the same building, and obtained and integrated two Hamilton® Microlab STAR liquid handling systems for automated accessioning and transfer of samples from collection tubes to 96-well plates prior to RNA extraction (see “EXCITE sample accessioning, RNA extraction, and RT-qPCR” section below).

Fourth, we obtained a CLIA extension from the existing Biochemical Genetics high-complexity CLIA laboratory in the Department of Pediatrics at UCSD, and established Standard Operating Procedures for each aspect of the workflow.

Finally, we built secure links between the INSPECT application and the UCSD instance of the EPIC electronic health record, as well as the California Reportable Disease Information Exchange (CalREDIE) Electronic Lab Reporting (ELR) system, using Redox as the integration application programming interface (API).

These changes were implemented in parallel, allowing us to translate the research workflow into a clinical assay in only forty days. The EXCITE lab was designed to accommodate up to 6,000 samples per day with next-day results.

### Validation of anterior nares samples

We clinically validated our clinical workflow using AN swabs collected in MTM (“ANM”), Mawi (“ANW”), and 5% SDS (“AND”). First, we performed a technical validation by determining the LOD of all three sample types using contrived samples with a range of SARS-CoV-2 viral particle (VP) concentrations, from 250 VP/mL up to 32,000 VP/mL, with each test performed in triplicate. We demonstrated that the sensitivity of our clinical workflow is excellent, with a technical LOD of 500 VP/mL for MTM and 250 VP/mL for Mawi and SDS (Table S2). Next, a second technical validation was performed using contrived samples containing 1,000 VP/mL; a minimum of 20 replicate samples were run per sample type, and a minimum of 19/20 were required to return a positive result via RT-qPCR (see “EXCITE sample accessioning, RNA extraction, and RT-qPCR” section below). All ANM and AND replicates returned a positive result, and 19/20 ANW replicates returned a positive result (Table S3). Last, we performed clinical validation on all three sample types, first validating ANM samples and then using the validated ANM as a comparator to clinically validate ANW and AND. We used remnant AN samples collected in MTM from 30 positive cases and 32 negative cases, kindly provided by Helix OpCo, LLC (San Diego, CA, USA). These samples had previously been tested/validated by Helix under their EUA (https://www.fda.gov/media/140420/download), and the test results obtained from Helix were used as the comparator for our results (Table S4). Following ANM validation and EUA authorization, we validated ANW and AND sample types with clinically-obtained positive and negative cases for SARS-CoV-2, and using the recently-validated ANM sample type as the comparator. Clinical validation required a minimum of 90% compliance between the EUA authorized comparator and the experimental sample types on the same samples. For ANW, 34 positives and 54 negatives were tested, and for AND, 38 positives and 55 negatives were tested (Table S4). The sensitivity (the ability of the test to correctly identify positives), specificity (the ability of the test to correctly identify negatives), and accuracy (overall probability that a test is correct) for each sample type can be seen in Table 1.

**Table 1.** Sensitivity, specificity, and accuracy of anterior nares swabs in MTM (ANM), Mawi (ANW), or 5% SDS (AND) media, tested for the presence of SARS-CoV-2 during clinical validation. Numbers in brackets are 95% confidence intervals.

|  | **Sensitivity** | **Specificity** | **Accuracy** |
| --- | --- | --- | --- |
| **ANM** | 100% (88.43 - 100) | 100% (89.11 - 100) | 100% (94.22 - 100) |
| **ANW** | 100% (89.11 - 100) | 100% (93.40 - 100) | 100% (95.80 - 100) |
| **AND** | 94.59% (81.81 - 99.34) | 100% (93.40 - 100) | 97.80% (92.29 - 99.73) |

### EXCITE sample collection

Self-swabbed anterior nares (AN) samples were collected using nylon flocked swabs (https://renonlab.en.made-in-china.com/product/zKEnIjuyEdrC/China-Medical-Virus-Test-Collection-Nylon-Flocked-Nasopharyngeal-Nasal-Swabs.html), which were placed in tubes containing 1 mL of either MTM, Mawi, or 5% SDS. All participants >12 years of age self-collected samples, and children 12 years of age or younger in participating schools had samples collected by trained staff at each school. Sample collection occurred on-site at participating fire departments and schools, and all samples were sent daily to the EXCITE lab for testing. For UCSD participants, sample test kits were available in dedicated vending machines throughout campus, with sample drop sites located next to the vending machines that were collected multiple times per day for testing by the EXCITE lab. EXCITE samples reported in this study were collected between September 15, 2020 and February 5, 2021.

### EXCITE sample accessioning, RNA extraction, and RT-qPCR

The workflow of the EXCITE lab was modeled after the SEARCH study, but updated and further automated to increase throughput. Sample accessioning was performed using Hamilton® Microlab STAR liquid handling systems. RNA was extracted using the MagMax Viral Pathogen (MVPII) kit, and extraction was semi-automated via the use of an epMotion® 5075 liquid handling workstation (Eppendorf) and a KingFisher™ Flex purification system (Thermo Fisher). A detailed protocol of the semi-automated sample accessioning and RNA extraction process used by the EXCITE lab is available on protocols.io (23).

Viral RNA was detected using the TaqPath COVID-19 Combo Kit (ThermoFisher Scientific, Waltham, MA, USA). RT-qPCR reaction preparation was performed using a Mosquito® HV Genomics robotic liquid handler (SPT LabTech), and miniaturized 3 µL RT-qPCR reactions were performed on a QuantStudio™ 7 Pro Real-Time PCR System (Applied Biosystems). A detailed protocol of the miniaturized and automated RT-qPCR process used by the EXCITE lab is available on protocols.io (22). Test results were determined as described above in the “SEARCH sample processing, RNA extraction, and RT-qPCR” section. Samples that tested positive through the EXCITE pipeline were re-extracted and re-amplified to confirm the positive result, and then reported to SD County. EXCITE participants received the results of every test they submitted, regardless of positivity.

### Data analysis and visualization

Data curation was conducted in R (version 4.0.3) using the ‘tidyverse’ (version 1.3.0) package (24) and the associated ‘readr’ (version 1.4.0) package. Statistical analyses including *t*- tests and Kruskall-Wallis tests were performed in R (version 4.0.3) or Python (version 3.6.11).

Chi-square tests and data summaries from Tables 5 and 6 were performed/created using SAS (version 9.4).

Data visualization was accomplished using a combination of R (version 4.0.3), Python (version 3.6.11), Tableau (Desktop version 2019.2.5 and Server version 2019.2.0), Excel 2010 (version 2103), Inkscape (version 1.0), and Leaflet (version 0.7.7). Figs 1C; S1; S2; S3; and S6 were created in Excel and edited in Inkscape. Figs 1A; 1B; 2; and 4 were created in Tableau and edited in Inkscape. Figs 3 and 5 were created in Python. Figs 6; 8; S4; S5; and S6 were created in R using the following packages: ‘lubridate’ (version 1.7.9.2) (25); ‘ggstatsplot’ (version 0.7.0) (26); ‘ggpubr’ (version 0.4.0); ‘tidyverse’ (version 1.3.0) (24) and the associated packages ‘readr’ (version 1.4.0), ‘readxl’ (version 1.3.1), ‘ggplot2’ (version 3.3.3), and ‘ggpubr’ (version 0.4.0). Figs 6 and 8 were created in Leaflet (https://leafletjs.com/) and OpenStreetMap (https://www.openstreetmap.org/copyright), using coordinate data from zipcodeR package (https://cran.r-project.org/web/packages/zipcodeR/index.html) and Google Maps (https://maps.google.com).

**Fig 1.**
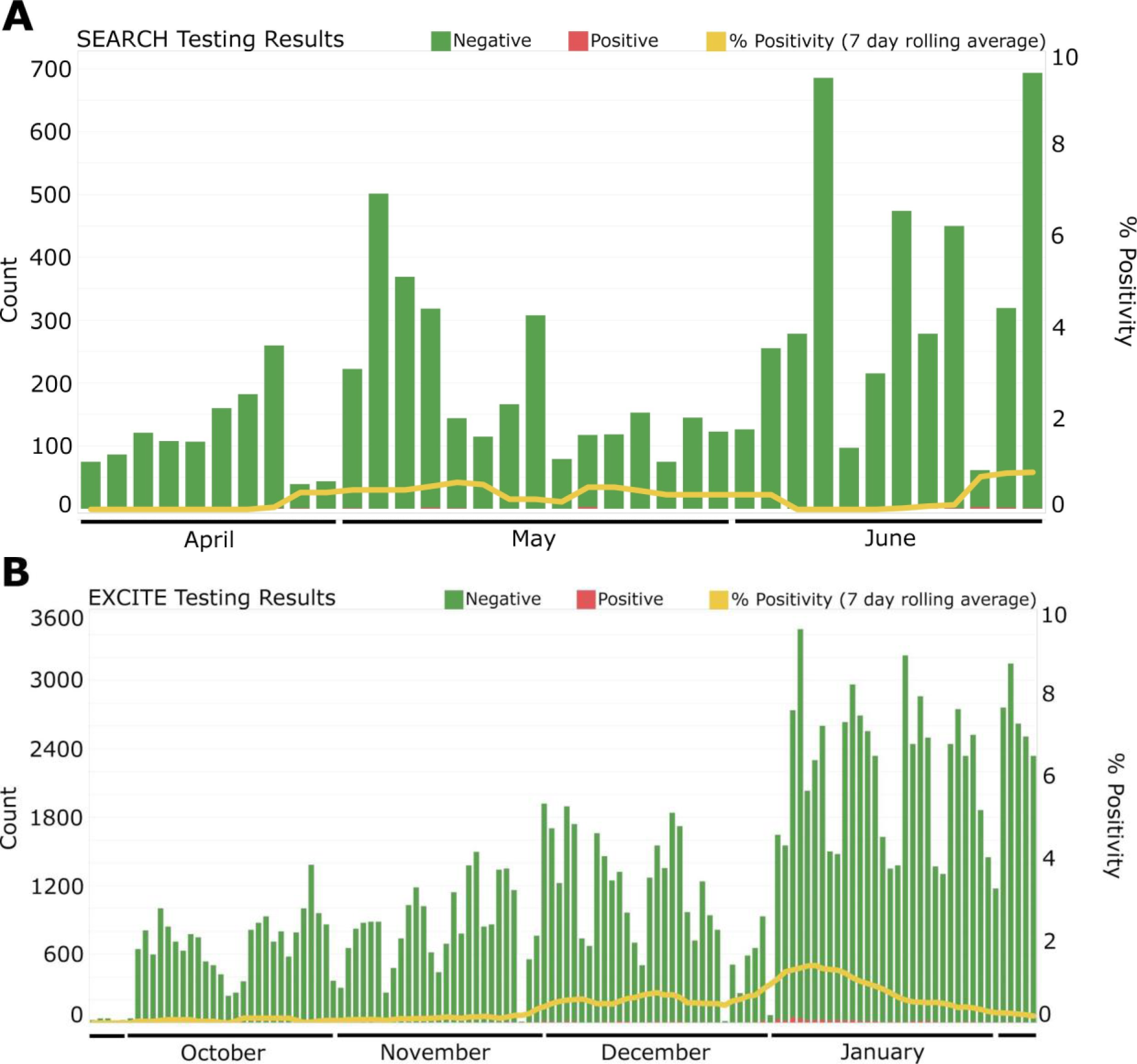
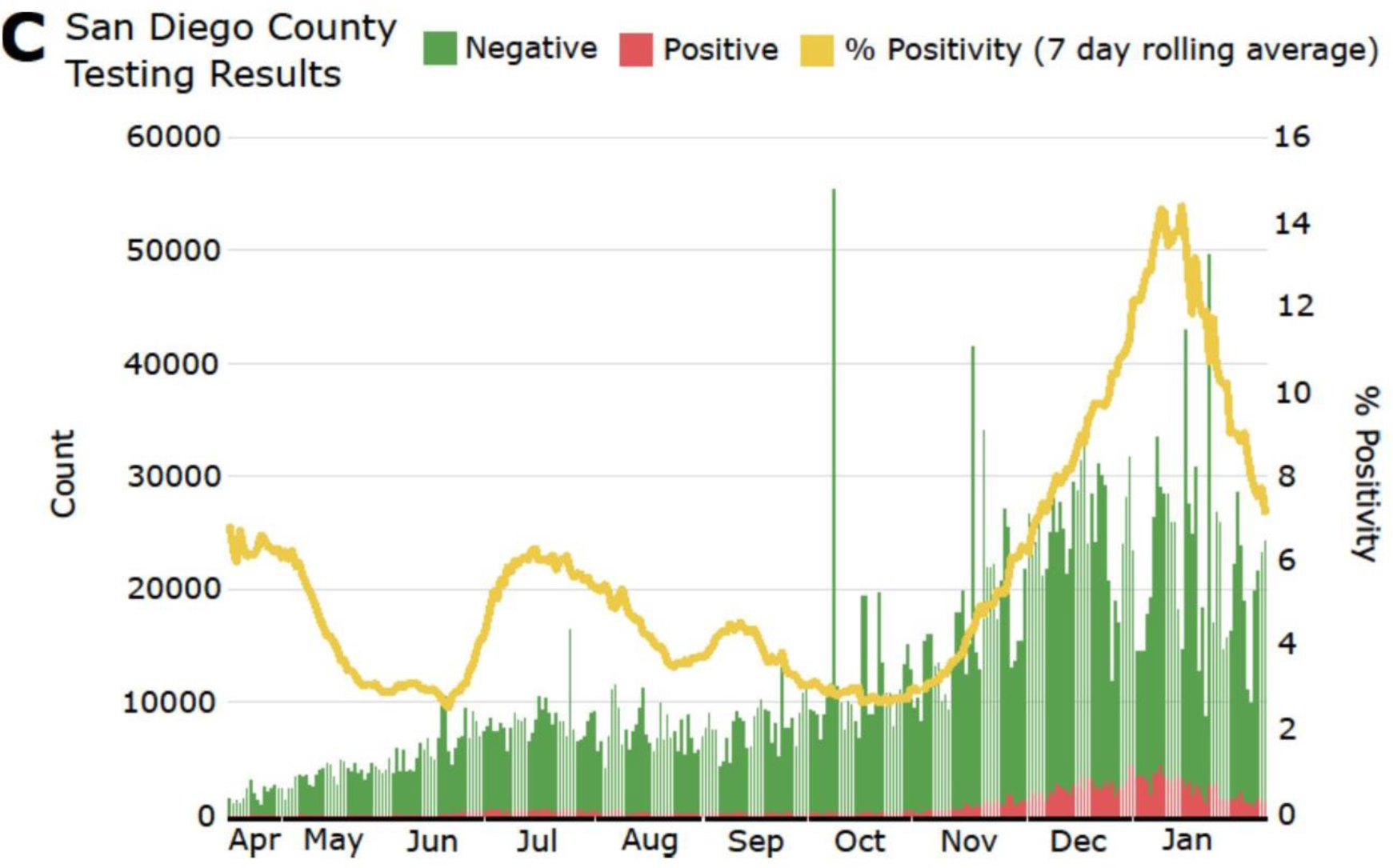
COVID-19 test results and rolling average of positivity rate for (A) SEARCH study participants (Apr 17 - Jun 30, 2020), (B) EXCITE lab participants (Sep 15, 2020 - Feb 5, 2021), and (C) San Diego County testing results (Apr 17, 2020 - Feb 5, 2021).

Full datasets, as well as scripts for data analysis and visualization used in this publication, are made available at https://osf.io/3fguz/.

## Results and Discussion

### Overall Testing Results

Samples were collected for the SEARCH study between April 17 and June 30, 2020, and by the EXCITE lab between September 15, 2020 and February 5, 2021. The SEARCH study was designed to screen healthcare workers and first responders from San Diego County (SD County); in particular, employees from Rady Children’s Hospital San Diego (RCHSD), Rady Children’s Institute for Genomic Medicine (RCIGM), Scripps Health, and the San Diego Fire-Rescue Department (SDFD) were screened, along with a small number of employees from other institutions who heard of the study by word of mouth and were allowed to participate. The EXCITE lab was designed for repeated screening of UCSD students and employees, but has also partnered with a number of private and public preschool-grade 12 schools and SDFD to provide repeated testing of their members. A recent study highlighted the potential positive implications of institutions expanding their COVID-19 testing efforts to include members of surrounding communities, which can benefit the institution itself as well as the community as a whole (27).

Both SEARCH and EXCITE included repeat testing of participants; in total, 8,066 nasopharyngeal (NP) swabs were obtained from 6,376 individuals via the SEARCH study, and 153,516 anterior nares (AN) swabs were obtained from 28,293 individuals via the EXCITE lab. During the time period of this study, the EXCITE lab processed an average of 1,124 samples per day. Additionally, the average total time from sample barcode scan to return of results was 15.2 ± 0.03 h, with an average of 6.6 ± 0.01 h of in-lab processing time (Fig S3). Results were returned within 24 h of receipt at the lab for 98.1% of samples, and between 24-60 h for 1.2% of samples. The latter were largely samples that returned with Inconclusive or Invalid results on the first analysis run, and were analyzed a second time before reporting a clinical result. We note that there was a small proportion (0.7%) of samples with processing times >60 h; these were primarily samples for which the lab did not initially receive the required patient-level demographic information to report a clinical result, and therefore needed to request additional information from the ordering healthcare provider prior to returning results.

The breakdown of test results by sample type is presented in Table 2. In the SEARCH study, 21 samples initially returned a positive result based on the RT-qPCR performed by the collaborating SEARCH research labs. Samples from all positive tests based on the research testing pipeline were sent to a CLIA-certified lab for validation using their clinical testing pipeline before results were returned to participants. Of the 21 initial positive tests, 18 were validated as true positives, and three were returned as false positives. Therefore, the false-positive rate for the SEARCH study was 0.037%. Because the EXCITE lab is itself a CLIA-certified lab, positive results were reported directly without external validation. Of the 752 true positive results identified through both SEARCH and EXCITE, 715 tests were obtained from unique individuals, with 35 individuals testing positive on two different occasions and one individual testing positive on three different occasions. The average time between positive tests for the same individual was 5 days; no individuals from this study were identified to be re-infected after recovering from COVID-19.

**Table 2.** SARS-CoV-2 RT-qPCR test results obtained from SEARCH and EXCITE, by sample type: AND (anterior nares swab in 5% SDS); ANM (anterior nares swab in MTM); ANW (anterior nares swab in Mawi); NPV (nasopharyngeal swab in VTM). All NPV samples were obtained from SEARCH, and all AND, ANM, and ANW samples were obtained from EXCITE. Samples where no viral target genes amplified but the internal control did amplify were considered negative. Samples where at least two out of three viral target genes amplified were considered positive. Samples where neither the viral target genes nor the internal control amplified were considered invalid. Samples where only one out of three viral target genes amplified were considered inconclusive.

|  | AND | ANM | ANW | NPV | Total |
| --- | --- | --- | --- | --- | --- |
|  | n (%) | n (%) | n (%) | n (%) | n (%) |
| Negative | 7288 (96.56) | 20959 (99.90) | 124182 (99.35) | 8032 (99.58) | 160461 (99.31) |
| Positive | 38 (0.50) | 15 (0.07) | 681 (0.54) | 18 (0.22) | 752 (0.47) |
| Invalid | 221 (2.93) | 3 (0.01) | 90 (0.07) | 12 (0.15) | 326 (0.20) |
| Inconclusive | 1 (0.01) | 2 (0.01) | 36 (0.03) | 4 (0.05) | 43 (0.03) |
| *Total* | *7548 (100)* | *20979 (100)* | *124989 (100)* | *8066 (100)* | *161582 (100)* |

While the SEARCH study only collected NP swabs in VTM, the EXCITE lab used AN swabs in three different preservation media; MTM, Mawi, and 5% SDS, corresponding to ANM, ANW, and AND sample types, respectively. All three media inactivate the SARS-CoV-2 virus, enabling self-collection. The ANM sample type was validated first, but was soon replaced by ANW and AND, which are non-toxic, allowing for unobserved self-collection. AND samples produced invalid results significantly more often than samples collected in other preservation media (Table 2); AND samples comprised only 4.7% of all samples, but 67.8% of all invalid test results. As a consequence, the EXCITE lab quickly discontinued the use of AND samples, focusing instead on ANW samples.

The percentage of positive tests from the SEARCH study was very low: only 17 participants (0.27%) had a positive COVID-19 test throughout the 2.5 month duration of the study (one of these participants submitted two positive samples, resulting in the count of 18 positive NPV test results in Table 2), and positive cases were spread relatively evenly throughout the timeframe of the study, with the rolling average always below 1% positivity (Fig 1A). Comparatively, SD County testing reported much higher positivity, with rolling average positivity ranging from ∼2-7% over the same time period (Fig 1C). In the EXCITE lab, positivity changed over time, starting low from mid-September through mid-November, and then rising following the Thanksgiving holiday (Nov 26, 2020), and rising again to a peak in early January 2021, after the winter holidays (Fig 1B). The number of tests performed per day increased throughout the fall, as the program was ramped up to test students residing in campus-owned housing or coming to campus every two weeks, and then decreased in late December 2020 when many students left the UCSD campus for the holidays. Overall testing was highest in early January 2021, when students residing in campus-owned housing or coming to campus were required to participate in intensive testing upon their return to campus (at days 1, 5, and 10 after return), and then went to a weekly testing cadence thereafter (see “Return to Learn at UCSD” section below for more details). The EXCITE data show a similar trend in positive test rates to the data from data from SD County (Fig 1C), except that the increase in positive cases in the fall started later (at the end of November 2020 for EXCITE, compared to the beginning of November for SD County) and show a slightly earlier drop in positive cases in January 2021 at EXCITE. The positivity rate of the SD County data was much higher than the rates reported by both SEARCH and EXCITE over the same time frame; the rolling average positivity rate for SEARCH never rose above 1%, and for EXCITE was less than 2% at its peak, as compared to the SD County data, where test positivity ranged from 2-15% across the same time period (April 17, 2020 - February 5, 2021). We note that the populations being tested by SD County are different from those tested in our study. Our data collection involved repeated screening of asymptomatic populations, while samples collected by SD County are more often from individuals who exhibit symptoms or have been exposed to a known COVID-19 case, likely explaining the large discrepancy in positivity rates. In particular, students at UCSD who were experiencing symptoms were referred to the Center for Advanced Laboratory Medicine (CALM), a separate CLIA-certified laboratory on campus, to be tested. Similarly, students and staff at the preschool-grade 12 partner schools were asked to stay home and obtain testing at SD County testing sites if they experienced any symptoms.

The winter holidays are typically a very social time for people living in the US, and despite the COVID-19 pandemic, millions of Americans traveled during the holidays in late 2020, contributing to a sharp increase in cases reflected both in our data and in the SD County data. The decline in UCSD cases throughout January 2021 was due to early detection of cases after return from winter break, minimal transmission on campus, and reduced community transmission in SD County. The decline in SD County cases observed in January 2021 is likely due to people returning to their homes following the winter holiday and the subsequent decrease in travel and social gatherings, and to a lesser extent, an increase in vaccination rates, which began in late 2020. Countries with higher vaccination rates in early 2021, including the UK and Israel, saw sharp declines in new COVID-19 cases and hospital admissions in February 2021, highlighting the remarkable efficacy of these vaccines, even after one dose (28, 29).

Almost all of the positive tests for this study came from the EXCITE lab dataset, and most positive tests were obtained between mid-November, 2020, and mid-January, 2021. UCSD participants made up most of the positive tests, which was expected because most of the EXCITE testing was conducted on UCSD students and staff (Fig 2). Only SDFD and UCSD participants were tested by both SEARCH and EXCITE (Fig 2). However, the UCSD population tested during the SEARCH study was different than that tested by the EXCITE lab. During the SEARCH study, some UCSD employees involved in the study chose to participate in testing.

**Fig 2.**
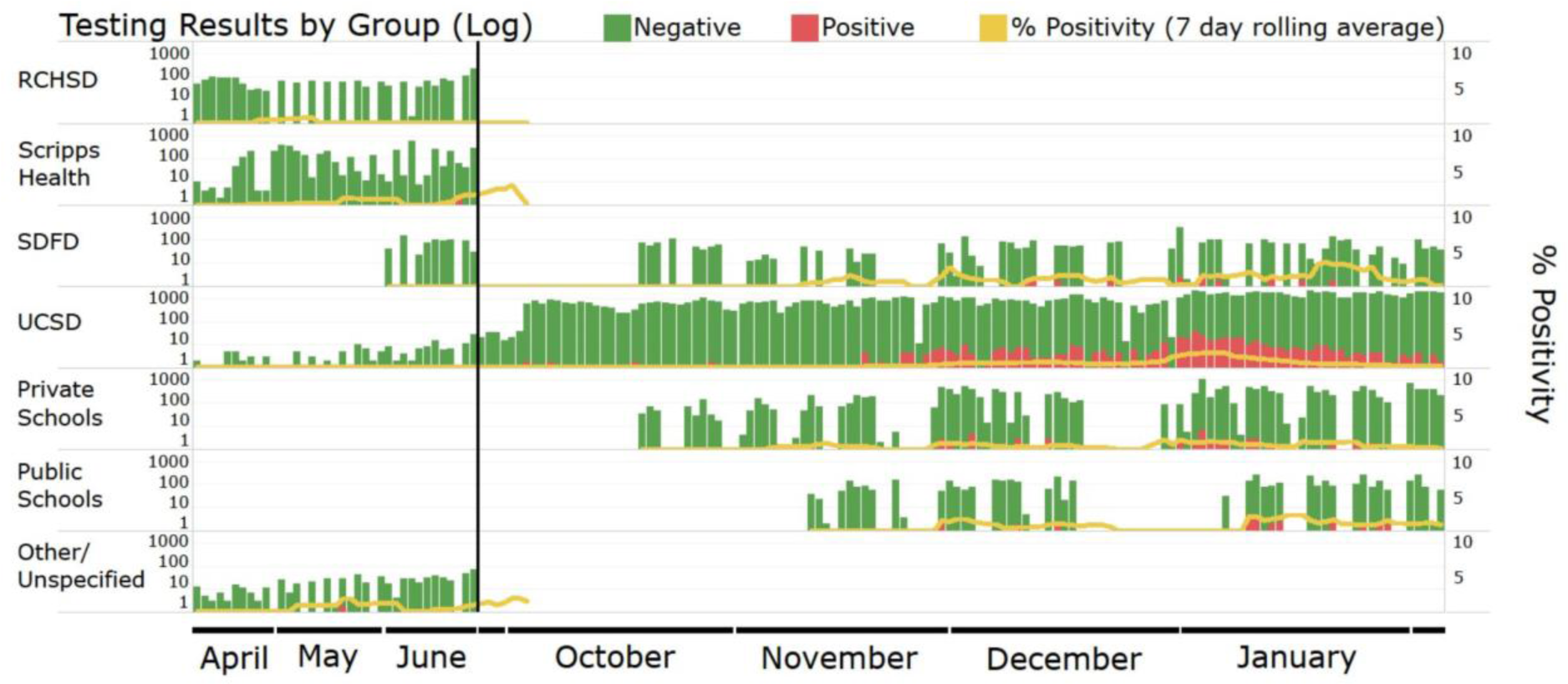
Log-transformed graph of COVID-19 testing results over time (by day), separated by source group. Participants being tested belonged to one of seven groups: RCHSD (Rady Children’s Hospital San Diego); Scripps Health (Scripps HealthCare); SDFD (San Diego Fire- Rescue Department); UCSD (University of California, San Diego); Private Schools (preschool- grade 12); Public Schools (preschool-grade 12); Other/Unspecified (healthcare workers from other locations).

While these employees likely also participated in testing through the EXCITE lab, they were not the main targets of either testing effort; the SEARCH study was designed to screen healthcare workers and first responders, and the EXCITE lab was established to screen students, faculty, and staff at UCSD and some partner institutions, with a focus on testing schools.

COVID-19 case data from SD County show that the 20-39 age group represents the highest number of cases (and case rates), followed by the 40-59 age group (Fig 3). The age groups represented by children (0-9 and 10-19) reported the lowest number of cases in the County. Within the EXCITE data from this study, participants in the 20-39 age group also made up the majority of cases, with all other age groups falling well below (Fig 3). This is largely due to the demographics of the EXCITE testing population, where the majority of participants were university students. We note that the 10-19 age range represents a higher proportion of cases in the EXCITE data than in the County data, also likely because of the large number of university students in the 18-19 age range in our study.

**Fig 3.**
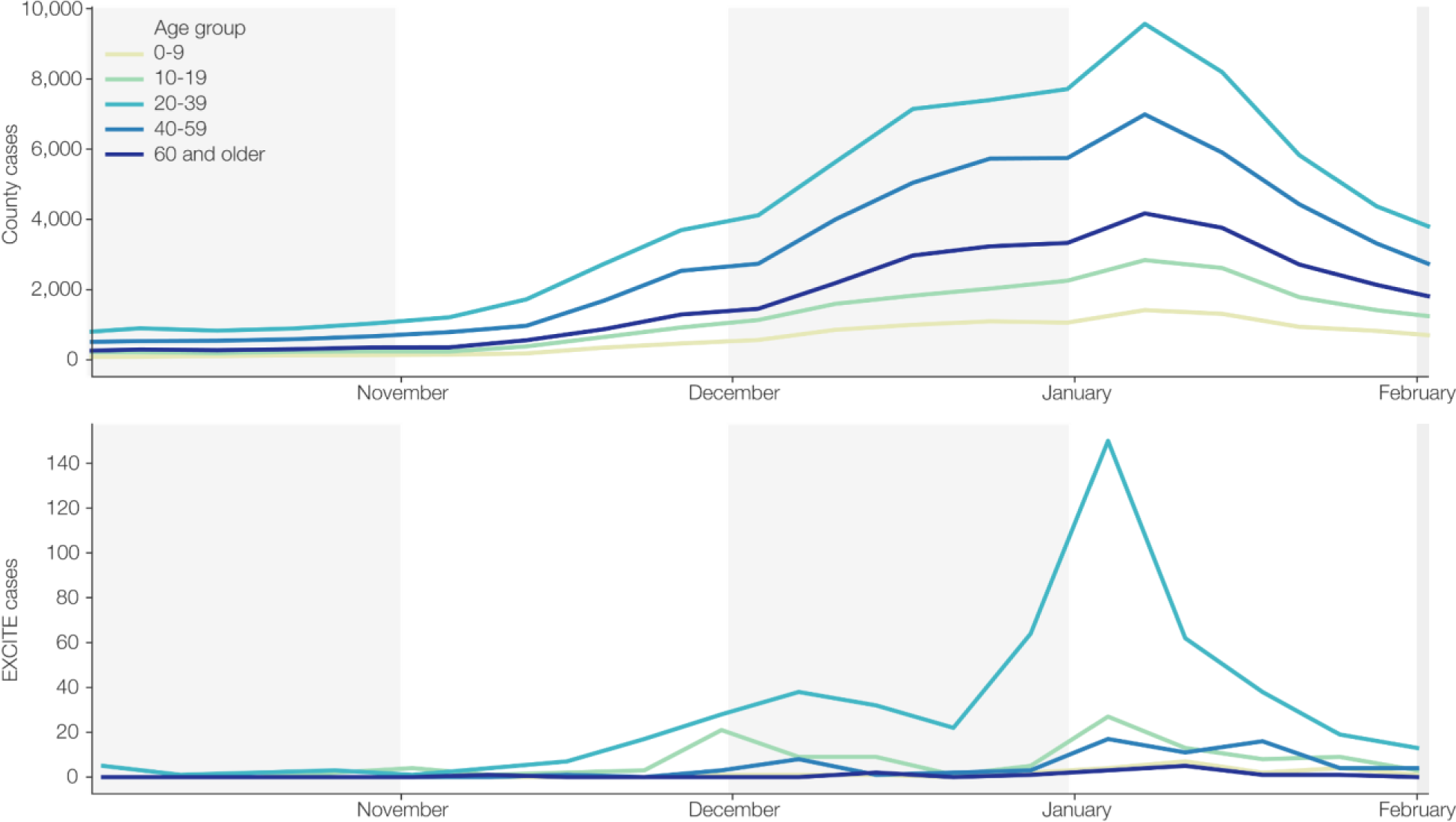
Number of positive cases per week, separated by age group. Top: San Diego County COVID-19 cases over time (Sep 15, 2020 to Feb 5, 2021). Bottom: EXCITE lab COVID-19 cases over the same timeframe.

No significant differences in viral load (estimated by Ct value) were observed for participants from different age groups (Fig S4). Previous studies have varied in their reports of viral loads in patients of different age groups. Some, like this current study, have not found any significant correlation between viral load and age groups (30–33), while others have (34–37). In the studies that did observe a difference, the results are conflicting: Euser et al. (37) reported that children <12 years had lower viral loads than adults, while Heald-Sargent et al. (36) reported children <5 years had higher viral loads than older children and adults. Additionally, Hasanoglu et al. (35) reported decreasing viral load with increasing age in adults, while To et al. (34) reported the opposite trend. Consistent with previous reports (10, 31), we did not find any significant difference in viral load between male and female participants (*t* = 0.02, df = 645.7, *p* = 0.98).

Only two of the three SARS-CoV-2 target genes were required to amplify for a test to be considered positive. The S-gene was the most common ‘dropout’ (i.e. the gene that failed to amplify), occurring in approximately 10% of positive cases; N-gene and ORF1ab dropouts occurred in approximately 1-2% of positive cases, respectively. With the exception of the cases in this study infected with the B.1.1.7 SARS-CoV-2 variant of concern, which show a characteristic S-gene dropout with low Ct values (<30) for the other two viral genes (38), samples with viral gene dropouts had higher average Ct values (i.e. lower viral load) as compared to samples without dropouts (Fig S5); samples containing gene dropouts had an average Ct value of 32.3 ± 1.1, while samples without dropouts averaged 23.0 ± 5.3 (*t* = -29.7, df = 2.2, *p* = 0.001). Seven cases were identified that were consistent with the B.1.1.7 variant (9.2% of all cases with S-gene dropouts), and the average Ct value for these seven cases was 21.9 ± 5.5.

### Symptom Reporting

Symptom reporting differed between SEARCH and EXCITE (Fig 4). In total, SEARCH participants filled out 7,489 symptom questionnaires (Table 3), and EXCITE participants filled out 18,318 (Table 4). Therefore, 92.8% of SEARCH samples were accompanied by symptom reporting, compared with just 11.9% of EXCITE samples. This discrepancy is due to the different nature of each screening system. The SEARCH study was designed as a prospective research study, so occupation and symptom information was requested from all participants.

**Table 3.** Symptom reporting by SEARCH participants who completed a symptom questionnaire at the time of testing. Results are per test, not per participant, in the case of participants testing on multiple occasions. Invalid and Inconclusive tests are included in the “All Tests” column but are not represented in subsequent columns.

| Symptoms | All Tests | Negative Tests | Positive Tests |
| --- | --- | --- | --- |
|  | n (%) | n (%) | n (%) |
| No symptoms | 6746 (90.08) | 6723 (90.17) | 10 (55.56) |
| Any symptoms | 743 (9.92) | 733 (9.83) | 8 (44.44) |
| Fever | 18 (0.24) | 15 (0.20) | 3 (16.67) |
| Chills | 47 (0.63) | 43 (0.58) | 4 (22.22) |
| Cough | 152 (2.03) | 147 (1.97) | 4 (22.22) |
| Sore throat | 177 (2.36) | 173 (2.32) | 4 (22.22) |
| Trouble breathing | 45 (0.60) | 44 (0.59) | 1 (5.56) |
| Stuffy nose | 205 (2.74) | 200 (2.68) | 5 (27.78) |
| Runny nose | 190 (2.54) | 185 (2.48) | 5 (27.78) |
| Nausea | 77 (1.03) | 74 (0.99) | 3 (16.67) |
| Vomiting | 13 (0.17) | 13 (0.17) | 0 (0) |
| Diarrhea | 61 (0.81) | 58 (0.78) | 3 (16.67) |
| Fatigue | 169 (2.26) | 165 (2.21) | 4 (22.22) |
| Myalgia | 149 (1.99) | 143 (1.92) | 5 (27.78) |
| Rash | 24 (0.32) | 24 (0.32) | 0 (0) |
| *Total* | *7489 (100)* | *7456 (100)* | *18 (100)* |

**Table 4.** Symptom reporting by EXCITE participants who completed a symptom questionnaire at the time of testing. Results are per test, not per participant, in the case of participants testing on multiple occasions.

| **Symptoms** | **All Tests** | **Negative Tests** | **Positive Tests** |
| --- | --- | --- | --- |
|  | **n (%)** | **n (%)** | **n (%)** |
| No symptoms | 18249 (99.62) | 18093 (99.68) | 90 (89.11) |
| Any symptoms | 69 (0.38) | 58 (0.32) | 11 (10.89) |
| Fever | 7 (0.04) | 2 (0.01) | 5 (4.95) |
| Cough | 11 (0.06) | 8 (0.04) | 3 (2.97) |
| Trouble breathing | 2 (0.01) | 2 (0.01) | 0 (0) |
| Fatigue | 10 (0.05) | 6 (0.03) | 4 (3.96) |
| Headache | 25 (0.14) | 20 (0.11) | 5 (4.95) |
| Anosmia | 4 (0.02) | 2 (0.01) | 2 (1.98) |
| Sore throat | 9 (0.05) | 8 (0.04) | 1 (0.99) |
| Stuffy/runny nose | 28 (0.15) | 23 (0.13) | 5 (4.95) |
| Nausea | 8 (0.04) | 8 (0.04) | 0 (0) |
| Diarrhea | 6 (0.03) | 6 (0.03) | 0 (0) |
| *Total* | *18318 (100)* | *18151 (100)* | *101 (100)* |

**Fig 4.**
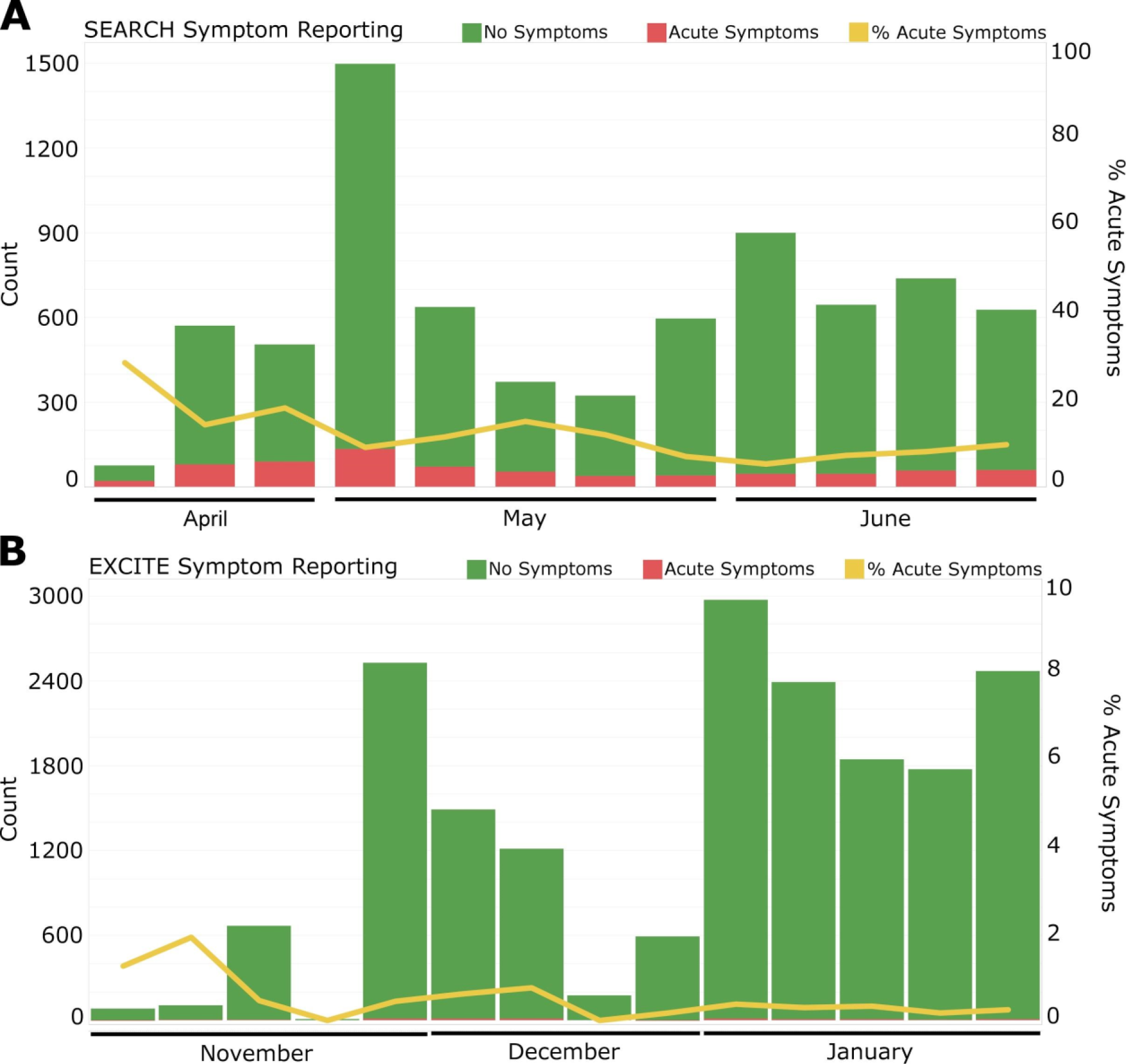
Symptom reporting by (A) SEARCH participants and (B) EXCITE participants over time.

The EXCITE lab was designed for high-throughput screening of asymptomatic populations, with no UCSD participants being asked to report symptoms at the time of testing, and participants from other partners being asked an abbreviated set of questions. Additionally, EXCITE participants were generally assumed to be asymptomatic, because any symptomatic individuals were encouraged to stay home and seek other means of testing. Conversely, at the time when SEARCH was implemented (April 2020), it was the only means of obtaining a COVID-19 test for many participants, resulting in a number of participants seeking out testing through SEARCH specifically because they were exhibiting symptoms they felt might be related to COVID-19. The timing of the SEARCH study also overlapped with the end of flu season, potentially explaining the higher number of participants reporting symptoms at the beginning of the study as compared to later in the study (Fig 4A). Indeed, when SEARCH participants waiting in line to be tested were asked why they chose to participate, their reasons mostly fell within four common themes: (1) early in the study, many believed their symptoms could represent COVID-19 illness but were unable to qualify for clinical testing because they were “not sick enough” per the testing triage protocols in place at the time; (2) later in the study, others believed their symptoms more likely represented seasonal allergies than COVID-19 but wanted to confirm, and understood the importance of reporting these symptoms nonetheless; (3) some reported intense stress regarding the pandemic and believed their symptoms of headache, fatigue, or muscle aches might be due to worry or poor sleep, but again wanted to confirm; (4) finally, several said they sought the privacy of testing outside their own healthcare workplaces. These unanticipated explanations for study participation illuminate some unmet needs for frontline workers in healthcare environments during the uncertainties of an unprecedented pandemic.

Among all SEARCH participants, the most common symptoms reported at the time of testing were stuffy nose, runny nose, sore throat, fatigue, and cough (Table 3). The least common symptoms reported were vomiting, trouble breathing, and fever. Among the participants who tested positive and also completed a symptom questionnaire, the most common symptoms at the time of testing were stuffy nose, runny nose, and myalgia (muscle pain), and the least common symptoms were rash, vomiting, and trouble breathing (Table 3). More than 90% of participants who tested negative reported no symptoms, compared with ∼55% of participants who tested positive; nearly 45% of participants who tested positive reported at least one symptom at the time of their test, with five being the average number of symptoms reported per person, out of a possible 14. The average age of SEARCH participants who completed symptom questionnaires was 42.9 ± 12.2. Of the 18 positive tests, 17 belonged to different individuals. One participant tested positive twice, reporting symptoms at the time of their first positive test, and reporting no symptoms at the time of their second positive test, nine days later. This participant tested negative on their third test, 16 days after their first positive test.

Among the EXCITE participants who completed symptom questionnaires, the most common symptoms reported at the time of testing were headache and stuffy/runny nose (Table 4). The least common symptoms reported were trouble breathing and anosmia (loss of sense of smell). Among the participants who tested positive and also completed a symptom questionnaire, the most common symptoms at the time of testing were fever, headache, and stuffy/runny nose, and the least common symptoms were trouble breathing, nausea, and diarrhea (Table 4). More than 99% of participants who tested negative reported no symptoms, compared with 89% of participants who tested positive; nearly 11% of participants who tested positive reported at least one symptom at the time of their test, with two being the average number of symptoms reported per person, out of a possible 10. The average age of EXCITE participants who completed symptom questionnaires was 24.1 ± 16.4.

Different symptom questions were asked of SEARCH and EXCITE participants. Some symptoms characteristic of COVID-19, such as anosmia (loss of sense of smell), were not included in the SEARCH study because sample collection began before this symptom was recognized as an indicator of COVID-19. Other symptoms, such as myalgia (muscle soreness) and rash, were included in the SEARCH questionnaire but not the EXCITE questionnaire. This is because only the most common cold, flu, and COVID-19 symptoms were included in the EXCITE questionnaire.

Of the 17 SEARCH patients who tested positive for SARS-CoV-2, 11 were available for regular telephone follow-ups, undertaken for 2-3 weeks in order to gain an understanding of symptom progression. Only three of these 11 people reported symptoms at the time of testing: one reported fever, one reported fatigue, muscle aches, and chills, and one reported cough, nasal congestion, and sore throat. Among the eight who did not report symptoms at the time of testing, five developed symptoms within 24 h of participating in the study, ranging from ageusia (loss of sense of taste) (*n* = 2) to sore throat with rhinorrhea (runny nose) (*n* = 1) to fever and cough (*n* = 2). One patient developed anosmia within 72 h, followed by severe fatigue and later a cough. Finally, two patients reported no symptoms of illness by two and three weeks, respectively. None of the ill participants required hospitalization, though one had an extended productive cough that began two days after testing positive and was placed on work leave for three weeks. While more than half of the SEARCH participants who tested positive for SARS-CoV-2 in our study did not report any symptoms at the time of testing, most went on to develop symptoms within 2-3 days of their first positive test, suggesting that within our study, being pre-symptomatic at the time of testing was more common than having an asymptomatic infection. Interestingly, shortness of breath/trouble breathing, which is considered a hallmark symptom of COVID-19, was reported by only one positive participant at the time of testing across both SEARCH and EXCITE, suggesting that this symptom may typically develop later on in the progression of the disease, as noted by some previous studies (39, 40), and may not be a good screening question to determine whether a person qualifies for a COVID-19 test. Reporting of the prevalence of shortness of breath/trouble breathing as a major symptom is inconsistent: one study also found shortness of breath to be a less common symptom (41), but other studies have reported shortness of breath to be one of the most common symptoms (42–44). However, it should be noted that these studies involved assessing symptoms in hospitalized patients, or even patients who suffered fatal cases of COVID-19, which represents a small fraction of infections.

Viral load was significantly correlated with the presence of symptoms for all three target genes (Fig 5), and only one of the 19 positive tests that were accompanied by a positive reporting of symptoms contained a gene dropout, suggesting that presentation of symptoms may indicate higher viral load in the body. Some previous studies have identified a correlation between viral load and symptom severity (45, 46), but most of these studies were conducted on hospitalized patients. Conversely, other studies have found viral loads to be unchanged (13) or even higher (35) in asymptomatic patients as compared to symptomatic patients. Interestingly, overall Ct values from SEARCH positives were higher than those from EXCITE positives, regardless of symptom reporting (*t* = -2.7, df = 18.1, *p* = 0.01) (Fig 5). It is possible that differences in the tested populations, sample type (nasopharyngeal versus anterior nares swabs), collection media, or RNA extraction kits used in SEARCH compared to EXCITE contributed to this observation.

**Fig 5.**
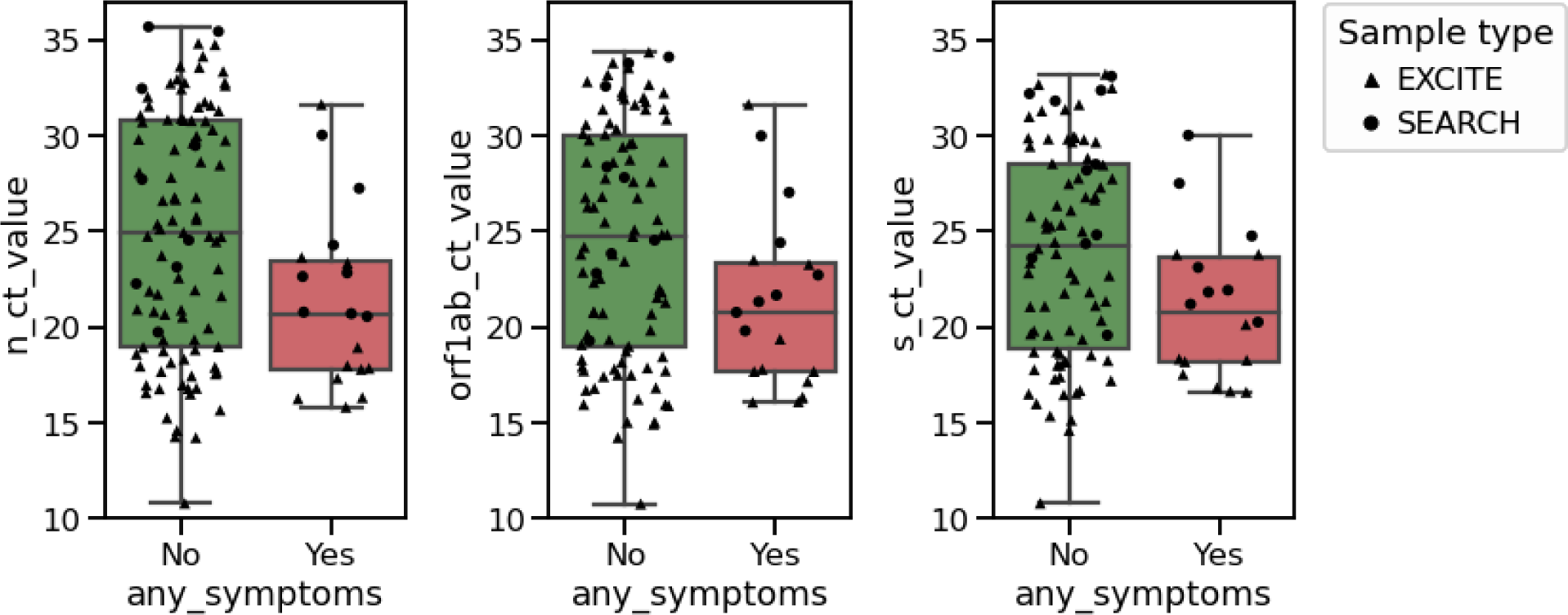
Ct values of three SARS-CoV-2 gene targets as a function of symptom reporting (*n* = 119). Samples from positive participants who also reported having symptoms had significantly lower Ct values (i.e. higher viral load) for all three gene targets, as compared with samples from positive participants who reported no symptoms (independent *t*-test *p*-value for N-gene, ORF1ab, and S-gene = 0.023, 0.031, 0.044, respectively).

### Healthcare Workers and First Responders

In order to keep the healthcare worker and first responder data together, we have included the SDFD first responders who were tested through the EXCITE lab in this section. Therefore, this section contains data from 6,786 individuals, while the SEARCH study itself only included 6,376 individuals. Of 11,964 total swabs, 3,935 were collected at RCHSD, 4,138 were collected at Scripps Health locations, and 3,891 were collected by the EXCITE lab. A proportion of study participants (1, 167) chose to be tested on multiple occasions, some as many as 22 times, although the majority of high-repeat participants were those from SDFD who were tested routinely through the EXCITE lab (Fig S6a). Of the 12,143 eligible Scripps employees who received an invitation to participate in this study, 3,838 (31.6%) participated at least once. Of the 1,470 eligible SDFD employees, 1,162 (79.0%) participated in testing at least once: 120 (8.2%) participated only in SEARCH, 410 (27.9%) participated only in EXCITE, and 632 (43.0%) participated in both. Approximately 8,000 Rady employees received an email inviting them to participate, but only approximately 6,000 opened the email. Additionally, this email was sent to employees of Rady Children’s Hospital San Diego (RCHSD), Rady Children’s Institute for Genomic Medicine (RCIGM), and Rady Children’s Specialists of San Diego (RCSSD), but on the intake form, only RCHSD was one of the pre-specified options, meaning employees of RCIGM and RCSSD likely would have selected the “Other” option as their employer, or simply not specified. A conservative estimate of participation, therefore, suggests that out of approximately 8,000 eligible Rady employees, 1,220 RCHSD employees (15.3%) participated. A more liberal estimate of participation would include both RCHSD and “Other” employees, and only those employees who opened their invitation email, resulting in 1,562 employees being tested out of an estimated possible 6,000 (26.0%). However, a small number of employees from Sharp HealthCare were informed of this study by word-of-mouth and were allowed to participate; these employees would likely have chosen “Other” or “Unspecified” as their organization as well. The true proportion of the target population tested is therefore likely somewhere between 15.3-26.0% for Rady employees. For simplicity, tables and Figs in this section combine the employees who selected “RCHSD” and “Other/Unspecified” into a single category, “Rady”. A small number of UCSD Health employees (112) also opted to be tested one or more times. These participants selected “UCSD” as their employer, but because UCSD employees were not targeted by the SEARCH study, we cannot estimate the proportion of employees tested for this population.

Participants were asked to add a job description on their intake forms. Because these job descriptions were free-text and not pre-specified categories, a wide range of occupations were reported. These occupations were manually combined into 14 major categories with the remainder being assigned to “Other/Unspecified” (Table 5). Approximately one-third (34.0%) of healthcare worker participants from Scripps and Rady were employed as nurses, with the next most common job category being general healthcare worker (24.2%), followed by doctor (11.9%). The proportions of nurses and doctors in this study is similar to their actual proportions at the hospitals and clinics from this study. The most common occupation among COVID-19 positive participants was first responder (66.7%) - this is because SDFD members additionally participated in repeated screening through the EXCITE lab in late 2020 and early 2021, and the entirety of the positive first responder cases came from that later time period, when cases in SD County were markedly higher. During the SEARCH study timeframe (April - June 2020), no SDFD members tested positive through our pipeline, and the most common occupations testing positive at that time were general healthcare workers and nurses (5 participants each). While some of these participants were presumed to have contracted the virus from work, others were able to trace their infection to other sources, such as a family gathering or a spouse with a public-facing job.

**Table 5.** Occupations of healthcare workers and first responders.

| **Occupation** | **All Participants**<br>**n (%)** | **Positive Participants**<br>**n (%)** |
| --- | --- | --- |
| Administrative | 302 (4.45) | 1 (1.96) |
| Dental worker | 23 (0.34) |  |
| Dentist | 30 (0.44) |  |
| Dietitian | 34 (0.50) |  |
| Doctor | 689 (10.15) | 1 (1.96) |
| First Responder | 1242 (18.30) | 34 (66.67) |
| Food Service | 37 (0.55) | 1 (1.96) |
| Healthcare worker (general) | 1340 (19.75) | 5 (9.80) |
| IT | 43 (0.63) |  |
| Management | 332 (4.89) | 2 (3.92) |
| Nurse | 1884 (27.76) | 5 (9.80) |
| Pharmacist | 76 (1.12) |  |
| Pharmacy worker | 29 (0.43) |  |
| Student | 35 (0.52) | 1 (1.96) |
| Other/Unspecified | 690 (10.17) | 1 (1.96) |
| *Total* | *6786 (100)* | *51 (100)* |

Demographic questions were asked of the participants; these questions were not mandatory for participation, but the majority of participants responded to most or all of the questions. The median age of healthcare workers was 41 for all target populations (Scripps Health, Rady, and SDFD) (Fig 6, Table 6). The median age of those participants who tested positive was significantly lower overall (35 years) and for healthcare workers (32 and 28 years for Scripps Health and Rady participants, respectively), but not significantly lower for SDFD participants (38.5 years) (Table 6). The overall data are consistent with CDC observations that the median age of COVID-19 infections has declined over time, from >40 years to <36 years between the months of March and July 2020 (47), and are in line with the median age (36 years) of confirmed COVID-19 cases among SD County residents at large (https://www.sandiegocounty.gov/content/dam/sdc/hhsa/programs/phs/Epidemiology/COVID-19%20Watch.pdf). Greene et al. (47) suggest that this decrease in the median age of confirmed COVID-19 cases is a result of changing testing patterns, not changes in the epidemiology of SARS-CoV-2 infection.

**Fig 6.**
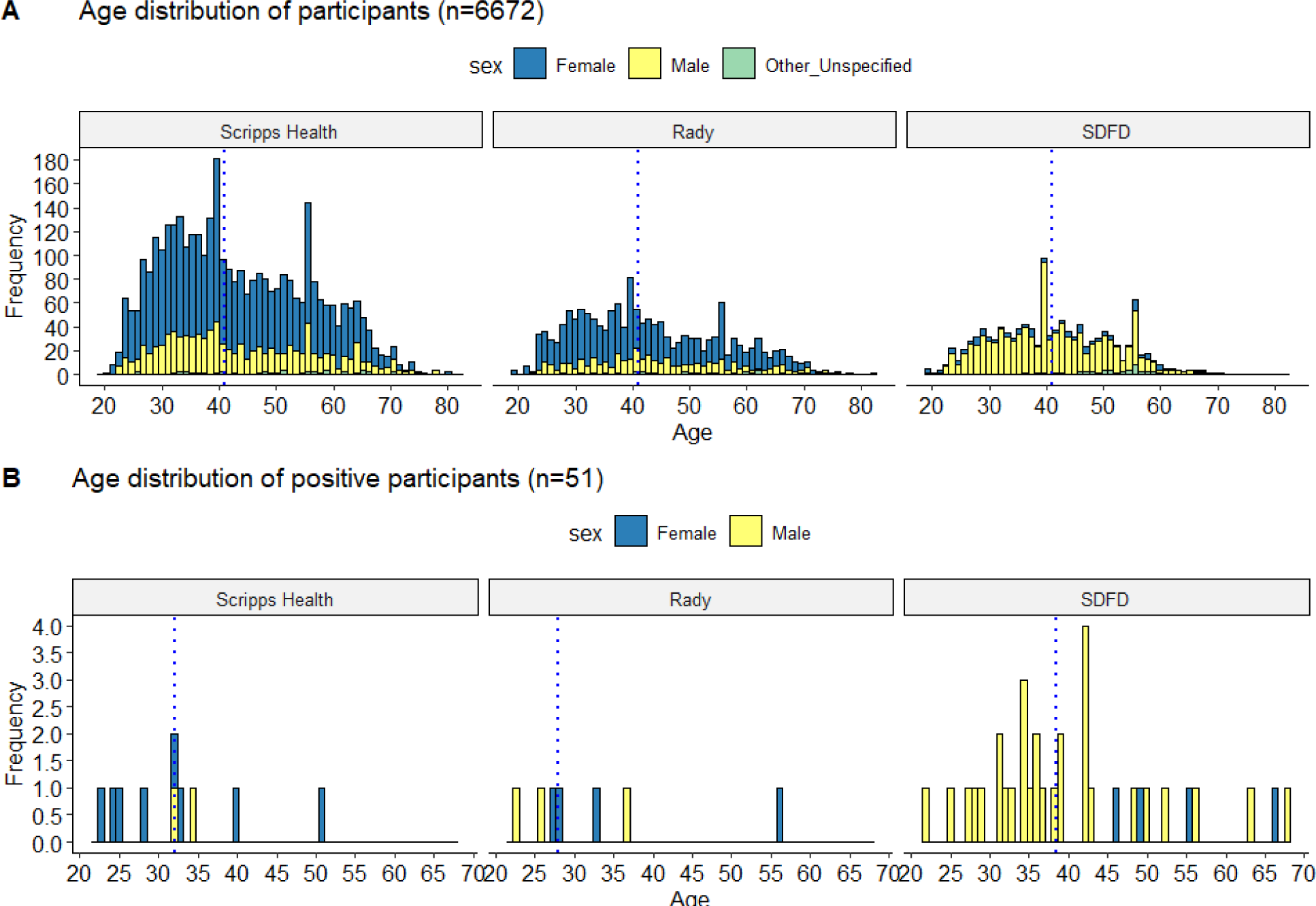
Age and sex distribution of (A) all participants and (B) positive participants from Scripps Health and Rady healthcare systems, and from the San Diego Fire-Rescue Department (SDFD). SEARCH participants who selected UCSD as their employer were excluded from this Fig because they were not a targeted population for the SEARCH study. The dotted line in each plot represents the median age.

Female participants made up 74.0% of healthcare workers, which is representative of the makeup of the workforce at the hospitals and clinics involved in this study, which are approximately 74% female across Scripps Health and approximately 83% female at RCHSD. Conversely, male participants made up 84.9% of first responders from SDFD, which is slightly lower than the national proportion of male firefighters (96%) (https://nfpa.org). Of the participants who tested positive for SARS-CoV-2, 70.6% of healthcare employees were female, while 85.7% of SDFD employees were male. Overall, males were significantly more likely to test positive than females (Table 6); however, when separated by source (Scripps Health, Rady, SDFD), no differences were observed. This is because the majority of the positive tests came from SDFD workers, who were predominantly male. Previous studies have suggested that males may be more susceptible to this virus (48), but our results are proportional to the populations tested in this study.

The majority of participants (72.1%) did not report their ethnicity, and almost one third (28.4%) did not report their race (Table 6). For the SEARCH study, race and ethnicity were presented as a single “check all that apply” question, which resulted in many people choosing only a race, only an ethnicity, or neither. Most participants did not select an ethnicity, and those that did select an ethnicity overwhelmingly selected Hispanic and did not also select a race. For the SDFD participants who were tested through the EXCITE lab, race and ethnicity were included as separate questions, which we believe improved reporting compliance. The EXCITE demographic data are presented for every SDFD participant who was tested through EXCITE, regardless of whether they were also originally tested through SEARCH. As a result, fewer SDFD participants did not specify their ethnicity or race, as compared to healthcare workers from Scripps Health and Rady (Table 6). SD County is ethnically diverse, with 34.1% of the population identifying as Hispanic (https://www.census.gov/). However, only 15.3% of the participants in this study selected Hispanic as their ethnicity. It is possible that the population in our study is not representative of the population of SD County, but it is also possible that Hispanic participants were less likely to report their ethnicity. There are many reasons why people choose not to divulge this information in their medical or employment records, or when participating in studies, including confusion over what category they fall into (especially when there are limited and pre-specified categories from which to choose) as well as fear of marginalization or receiving unequal quality of care (49). Additionally, both race and ethnicity having been combined into a single “check all that apply” question for the SEARCH study likely caused additional confusion. Most people who selected both a race and an ethnicity identified as White and Hispanic. In the case of the SEARCH study, it may not have been clear to participants that they were meant to select more than one option, and therefore most participants selected the one answer they identified most strongly with.

**Table 6.** Demographic information of SEARCH healthcare worker participants (Scripps Health, Rady) and the San Diego Fire- Rescue Department (SDFD) first responders. Participants who selected UCSD as their employer are included in the Overall category. The Positive column includes individuals who tested positive for COVID-19 at least once, while the Negative column includes individuals who never tested positive. Chi-square and *t*-tests were performed to test for differences among groups. Differences were considered significant at p ≤ 0.05 and are indicated in bold. The first column (Total) was not included in statistical analysis and was included to show the demographics of the entire population, regardless of test result.

|  | Total | Overall |  |  | Scripps |  |  | Rady |  |  | SDFR |  |  |
| --- | --- | --- | --- | --- | --- | --- | --- | --- | --- | --- | --- | --- | --- |
|  |  | Positive | Negative | *p*-value | Positive | Negative | *p*-value | Positive | Negative | *p*-value | Positive | Negative | *p*-value |
| Age, mean (SD), median | 42.7 (12.1)<br>41 | 38.0 (11.4)<br>35 | 42.7 (12.1)<br>41 | **0.005** | 32.2 (8.4)<br>32 | 43.3 (12.4)<br>41 | **0.005** | 32.9 (11.2)<br>28 | 43.1 (12.5)<br>41 | **0.03** | 40.7 (11.5)<br>38.5 | 41.2 (10.2)<br>41 | 0.79 |
| Age, n (%) |  |  |  |  |  |  |  |  |  |  |  |  |  |
| < 30 | 947 (14.6) | 13 (1.4) | 934 (98.6) | 0.07 | 4 (0.8) | 494 (99.2) | 0.15 | **4 (1.7)** | 229 (98.3) | **0.04** | 5 (2.9) | 165 (97.1) | 0.14 |
| 30-39 | 2027 (31.3) | 20 (1.0) | 2007 (99.0) |  | 4 (0.3) | 1156 (99.7) |  | 2 (0.4) | 471 (99.6) |  | 14 (3.9) | 345 (96.1) |  |
| 40-49 | 1562 (24.2) | 9 (0.6) | 1553 (99.4) |  | 1 (0.1) | 815 (99.9) |  | 0 (0.0) | 381 (100.0) |  | 8 (2.3) | 345 (97.7) |  |
| 50-59 | 1224 (18.9) | 6 (0.5) | 1218 (99.5) |  | 1 (0.1) | 702 (99.9) |  | 1 (0.4) | 271 (99.6) |  | 4 (1.7) | 235 (98.3) |  |
| 60+ | 707 (10.9) | 3 (0.4) | 704 (99.6) |  | 0 (0.0) | 466 (100.0) |  | 0 (0.0) | 204 (100.0) |  | 3 (8.6) | 32 (91.4) |  |
| Race, n (%) |  |  |  |  |  |  |  |  |  |  |  |  |  |
| Unspecified | 1926 (28.4) | 10 (0.5) | 1916 (99.5) | **0.03** | 6 (0.5) | 1183 (99.5) | 0.19 | 4 (0.7) | 586 (99.3) | 0.70 | 0 (0.0) | 114 (100.0) | 0.06 |
| White | 3432 (50.6) | **30 (0.9)** | 3402 (99.1) |  | 1 (0.1) | 1719 (99.9) |  | 3 (0.4) | 795 (99.6) |  | 26 (3.1) | 821 (96.9) |  |
| Black/AA | 173 (2.6) | **4 (2.3)** | 169 (97.7) |  | 0 (0.0) | 102 (100.0) |  | 0 (0.0) | 32 (100.0) |  | 4 (10.3) | 35 (89.7) |  |
| Other/mixed | 210 (3.1) | **4 (1.9)** | 206 (98.1) |  | 0 (0.0) | 77 (100.0) |  | 0 (0.0) | 27 (100.0) |  | 4 (3.9) | 100 (96.1) |  |
| Asian | 892 (13.1) | 3 (0.3) | 889 (99.7) |  | 3 (0.5) | 647 (99.5) |  | 0 (0.0) | 199 (100.0) |  | 0 (0.0) | 31 (100.0) |  |
| Indigenous/NA | 20 (0.3) | 0 (0.0) | 20 (100.0) |  | 0 (0.0) | 13 (100.0) |  | 0 (0.0) | 4 (100.0) |  | 0 (0.0) | 3 (100.0) |  |
| Pacific Islander | 133 (2.0) | 0 (0.0) | 133 (100.0) |  | 0 (0.0) | 88 (100.0) |  | 0 (0.0) | 21 (100.0) |  | 0 (0.0) | 24 (100.0) |  |
| Ethnicity, n (%) |  |  |  |  |  |  |  |  |  |  |  |  |  |
| Unspecified | 4891 (72.1) | 11 (0.2) | 4880 (99.8) | **< 0.0001** | 6 (0.2) | 3254 (99.8) | 0.10 | 5 (0.4) | 1361 (99.6) | 0.39 | 0 (0.0) | 169 (100.0) | **0.02** |
| Hispanic | 1036 (15.3) | **11 (1.1)** | 1025 (98.9) |  | 4 (0.7) | 554 (99.3) |  | 2 (0.7) | 294 (99.3) |  | 5 (3.0) | 161 (97.0) |  |
| Non-Hispanic | 859 (12.7) | **29 (3.4)** | 830 (96.6) |  | 0 (0.0) | 21 (100.0) |  | 0 (0.0) | 9 (100.0) |  | **29 (3.5)** | 798 (96.5) |  |
| Sex, n (%) |  |  |  |  |  |  |  |  |  |  |  |  |  |
| Female | 4270 (62.9) | 16 (0.4) | 4254 (99.6) | **< 0.0001** | 8 (0.3) | 2822 (99.7) | 1.00 | 4 (0.3) | 1244 (99.7) | 0.47 | 4 (3.0) | 129 (97.0) | 0.78 |
| Male | 2373 (35.0) | **35 (1.5)** | 2338 (98.5) |  | 2 (0.2) | 950 (99.8) |  | 3 (0.8) | 380 (99.2) |  | 30 (3.0) | 957 (97.0) |  |
| Other/unspecified | 143 (2.1) | 0 (0.0) | 143 (100.0) |  | 0 (0.0) | 57 (100.0) |  | 0 (0.0) | 40 (100.0) |  | 0 (0.0) | 42 (100.0) |  |

More healthcare workers identified as Asian (16.9% for Scripps Health and 11.9% for Rady, as compared to 2.7% for SDFD), while more SDFD participants identified as Other/Mixed Race (9.0% for SDFD, compared to 2.0% for Scripps Health and 1.6% for Rady). In all three populations, the majority of participants identified as White (44.8%, 47.7%, and 72.9% for Scripps Health, Rady, and SDFD, respectively). However, direct comparisons between healthcare workers and SDFD first responders may be inappropriate, since the methods used to obtain demographic information were different for these populations. Overall, participants identifying as White, Black/African American, and Other/Mixed Race were more likely to test positive (*p* = 0.03), but no differences were observed when separated by source (Table 6). Non- Hispanic SDFD participants were more likely to test positive than Hispanic or Unspecified participants (*p* = 0.02), but no similar differences were observed among healthcare workers.

Differences were considered significant at *p* ≤ 0.05 and are indicated in bold. The first column (Total) was not included in statistical analysis and was included to show the demographics of the entire population, regardless of test result. Participant-provided zip codes of residence were overlaid on a map of SD County (Fig 7). Study participants were spread throughout the County, with the highest concentration in San Diego and the communities slightly further north towards La Jolla, which was expected given that many of the healthcare systems monitored in this study are located in or near these regions. However, the broad distribution of participants across SD County suggests that the mobile testing sites were successful in enabling a more representative population to participate. The distribution of positive participants largely matched the overall distribution of participants, with concentrations in more heavily tested/populated areas. We note that even when participants chose to be tested multiple times, they are represented only once in Fig 7.

**Fig 7.**
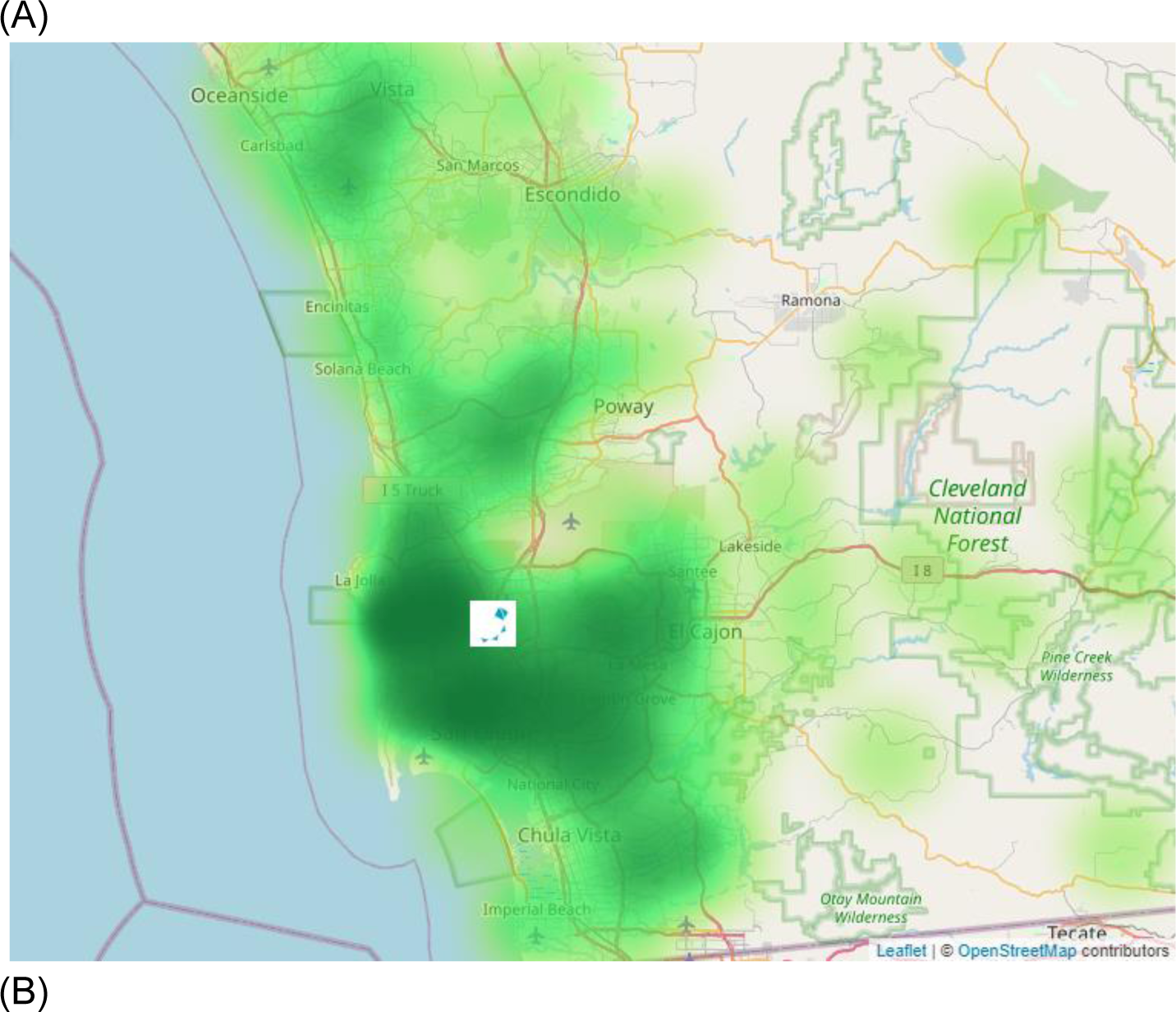
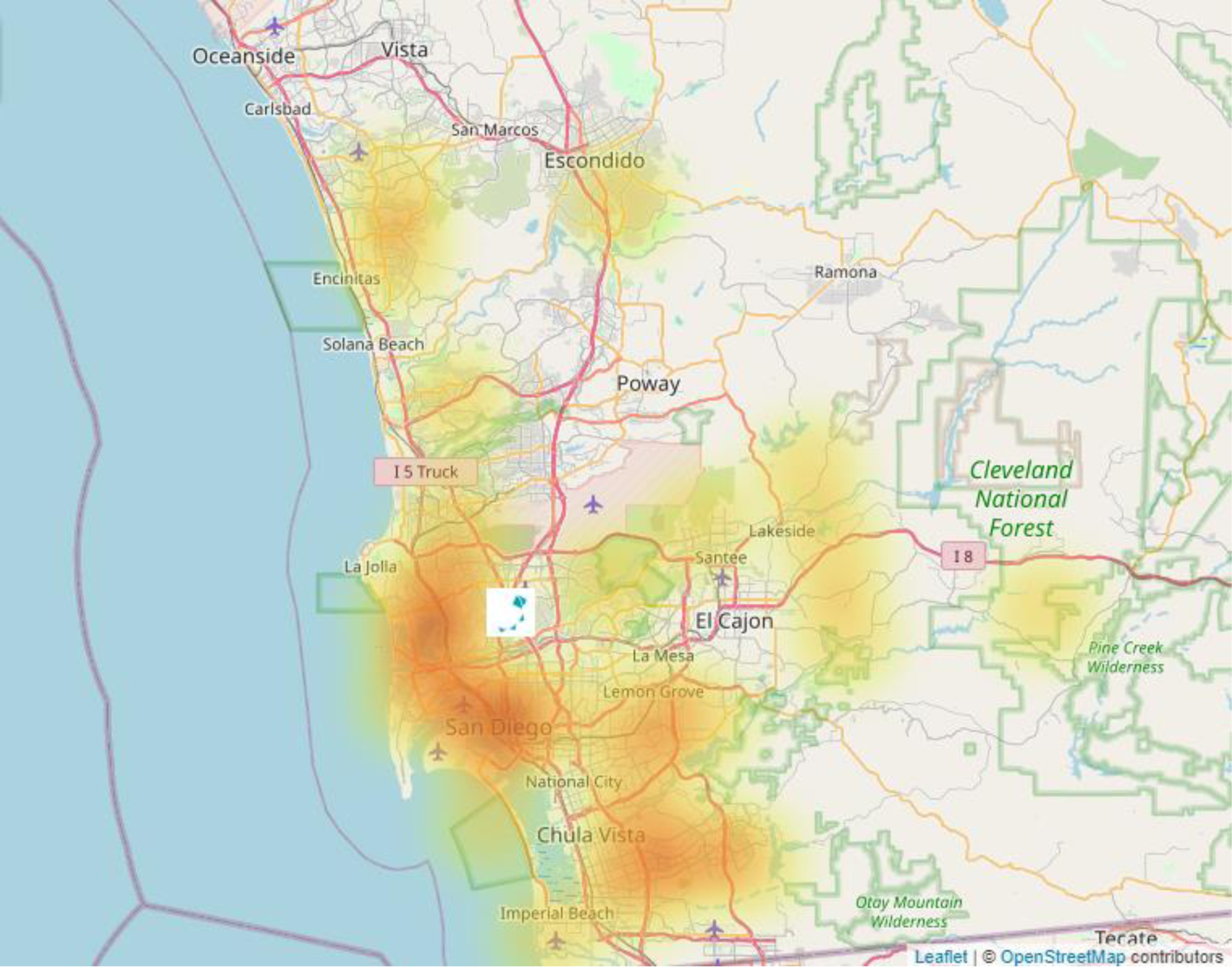
Zip codes of residence of (A) all healthcare worker and first responder participants tested through the SEARCH study and EXCITE lab, and (B) all healthcare worker and first responder participants who tested positive for COVID-19. The deepness of color is proportionate to the number of participants who chose that zip code as their area of residence. The icon on both images represents the location of Rady Children’s Hospital San Diego, where the permanent testing location was established during the SEARCH study.

### UCSD and Preschool-Grade 12 Schools

#### Population Demographics

Data from UCSD, 11 private preschool-grade 12 (P-12) schools, and a group of 13 public P-12 schools located in SD County are included in this section. All participants were tested via the EXCITE lab at UCSD, and this section contains data from 27,252 individuals, with 21,221 coming from UCSD, 1,281 coming from public P-12 schools, and 4,750 coming from private P-12 schools. Because EXCITE was designed for repeated testing of asymptomatic populations, the majority of participants (81.7%) were tested on multiple occasions, some over 40 times (Fig S6b). The participants who tested most frequently were likely a combination of EXCITE lab employees and UCSD student athletes, who were required to test frequently to decrease the chance that their activities would be interrupted due to COVID-19 infections.

Demographic information was obtained from the participants. In this section, each participant was counted once, regardless of how many times they participated in testing; for positivity status, “positive” participants are those who tested positive through EXCITE at least once, while “negative” participants are those who never tested positive. For UCSD participants, demographic information was gathered from pre-existing student/employee records. For P-12 school participants, demographic information was gathered either as part of the consent process for participation in this testing program, from student/employee records, or a combination of the two. The median age was 22 for UCSD participants, 11 for public school participants, and 16 for private school participants (Fig 8, Table 7). Most participants from each group belonged to the ‘student’ age ranges appropriate for each educational facility. Overall, participants were evenly split between male and female; however, within the P-12 schools, sex was evenly split for student age ranges but skewed female for adult (teachers and staff) age ranges (Fig 8). We found no differences in positivity rate by sex, overall or within any of the three populations studied. Our finding was similar to the overall case reporting by sex by SD County (https://www.sandiegocounty.gov/). However, SD County did report a higher fatality rate for males than for females, and previous research investigating sex disparities also found that while males are not more likely to test positive, they are more likely to be hospitalized or die from COVID-19 (50). We have no information on whether any of the participants testing positive through the EXCITE lab were hospitalized.

**Fig 8.**
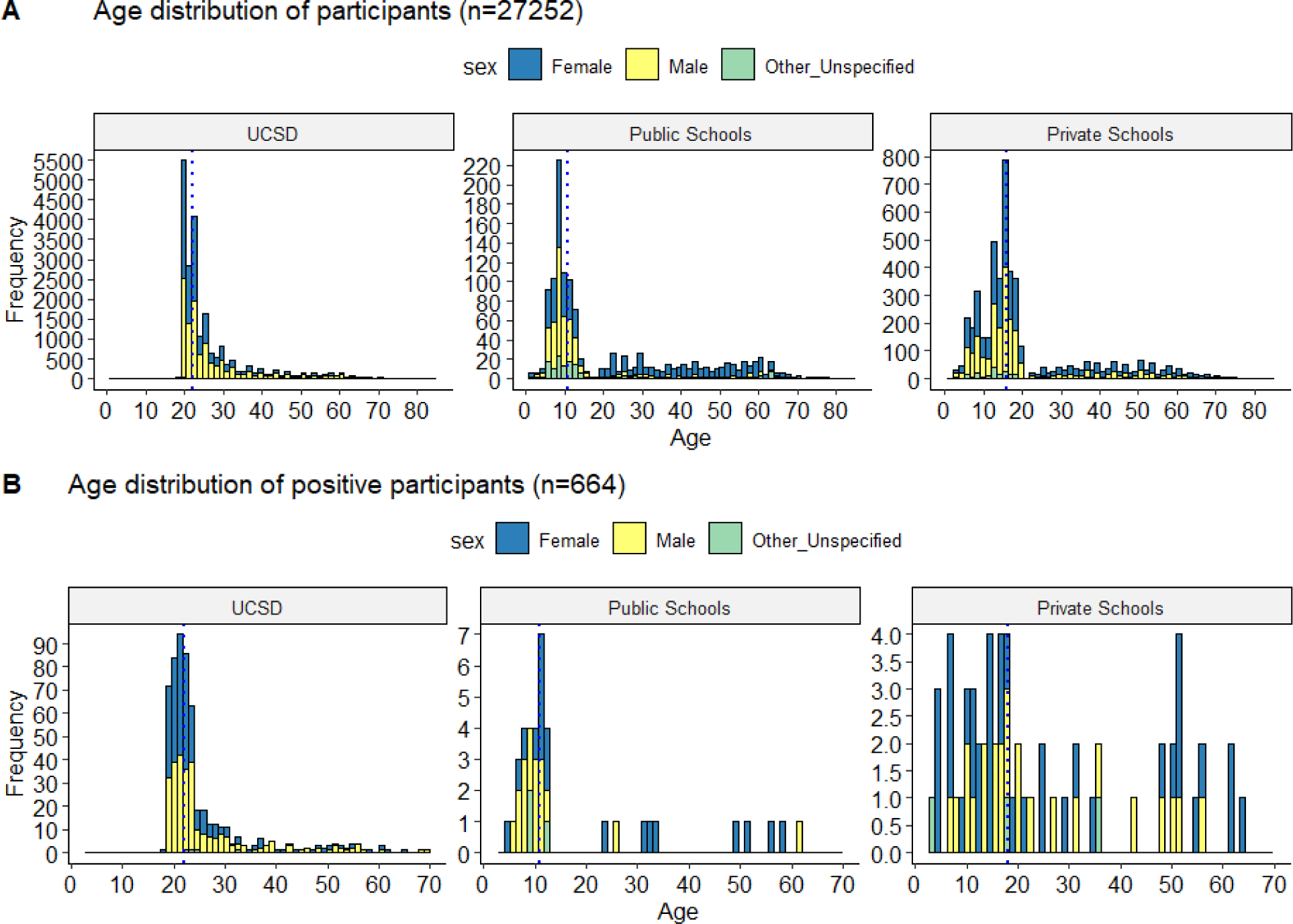
Age and sex distribution of (A) all EXCITE UCSD/school participants, and (B) all COVID- 19 positive participants from the same population. The dotted line in each plot represents the median age.

**Table 7.**
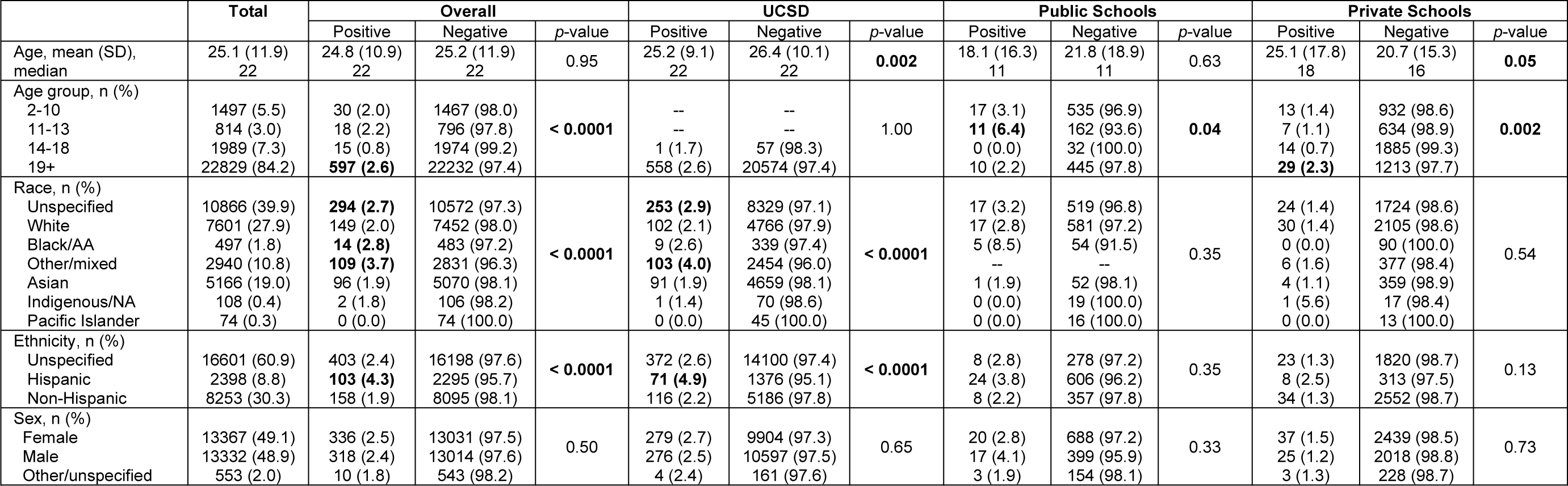
Demographic information of EXCITE participants. The Positive column includes individuals who tested positive for COVID- 19 at least once, while the Negative column includes individuals who never tested positive. Chi-square and *t*-tests were performed to test for differences among groups. Differences were considered significant at *p* ≤ 0.05 and are indicated in bold. The first column (Total) was not included in statistical analysis and was included to show the demographics of the entire population, regardless of test result.

Adults 19+ were overall more likely to test positive than younger age groups (*p* < 0.0001), partly because the majority of the participants were adults from UCSD (Table 7). While almost all UCSD participants were aged 19+, the average age of people testing positive at UCSD was slightly lower than those testing negative (25.2 vs. 26.4, respectively; *p* = 0.002). For private school participants, we found that students tested positive less frequently than adults, but this trend was not extended to public school participants. At private schools, adults were more likely to test positive than children (2.3% of adults as compared to 1.0% of children, *p* = 0.002), but at public schools, children aged 11-13 were the most likely to test positive (*p* = 0.04). This could be due to differences in testing uptake by the schools: for most private schools, students and staff who attended in-person learning at school were required to participate in regular testing, but for public schools, participation was voluntary. This testing program was designed for asymptomatic screening, and participants who felt sick were discouraged from coming to school and were encouraged to seek testing from SD County testing sites or primary care providers. Therefore, these data do not show the complete picture, but they do show that with proper protective measures, students attending in-person schooling are not testing positive more frequently than the general population (Fig 3), and in the case of the private schools in this study, may be testing positive less frequently than adults in the same settings. Our private school results are similar to other studies, which have also shown that transmission rates in schools are low (5, 6), and that students were less likely to test positive than staff in educational settings (8). Also of note, there were no outbreaks in participating schools in this study that were attributable to in-school transmission. The low positivity among school-aged children provides an argument for opening P-12 schools for in-person learning; with teachers and some school-aged children eligible for vaccination across the US, and children representing the age group that is currently least likely to test positive, our study and others suggest that schools can re- open with minimal risk of community transmission (5–8). However, it should be noted that the low rate of positivity among children observed at the County level (Fig 2) could be a result of school closures during this time, which would have drastically reduced the number of contacts each child would have.

A large proportion of participants chose not to identify their race (39.9%) or ethnicity (60.9%) (Table 7). Across all three populations studied, approximately 40% of participants did not report their race, but the reporting of ethnicity varied by population: two-thirds (67%) of UCSD participants did not report their ethnicity, compared to ∼40% of private school participants and just ∼20% of public school participants. Because UCSD student/employee records default to unspecified race and ethnicity unless a participant changes their status themselves, or their status is changed by a healthcare worker during a medical visit, this could account for a significant proportion of the participants who did not report a race or ethnicity. However, the discrepancy between race reporting and ethnicity reporting at UCSD suggests that there is a portion of this population reporting race, but declining to report ethnicity. Similar proportions of participants did not report their race at both public and private schools, but twice as many private school participants (proportionally) declined to report their ethnicity. As with the SEARCH study, legal status in the US was not a requirement for participation in the EXCITE testing program, nor was it a question asked of potential participants.

More UCSD participants identified as Asian (22.4%, as compared to 7.6% and 4.1% for private and public schools, respectively), while more P-12 school participants identified as White (44.9% and 46.7% for private and public schools, respectively, as compared to 22.9% for UCSD). While the overall number of participants identifying as Black/African American, Indigenous/Native American, or Pacific Islander was comparatively low, more public school participants selected these options as their race identity (Table 7). Additionally, approximately half of public school participants identified as Hispanic, compared to just 6.8% of UCSD and private school participants (Table 7). Overall, participants identifying as Black/African American or Other/Mixed Race were more likely to have a positive test, as well as those with an unspecified race (*p* < 0.0001), although when separated by group the result was not statistically significant for P-12 schools. Additionally, participants identifying as Hispanic were also more likely to test positive, although this difference was only statistically significant for UCSD participants (*p* < 0.0001), not for P-12 schools. Even though the result was not statistically significant, there was a trend toward a larger proportion of Black/African American and Hispanic participants from the public schools testing positive for COVID-19 (Table 7). Similar racial and ethnic disparities have been noted previously by other researchers, highlighting the disproportionate toll the COVID-19 pandemic has had on already-marginalized population in the US (51–55). In particular, Gil et al. (51) identified potential reasons why Hispanic people may test positive for COVID-19 more frequently, including: higher rates of coexisting medical conditions; lower rates of health insurance; immigration status; and language barriers. Similarly, Millett et al. (54) note that counties with a larger Black population were also more likely to have higher rates of air pollution, comorbidities, and lower rates of health insurance. Factors such as language barriers and lack of health insurance prevent access to testing or hospital care, while factors such as air pollution and comorbidities can increase the severity of an infection.

Participant-provided zip codes of residence were overlaid on a map of SD County (Fig 9). Participants involved in testing through the EXCITE lab came from all across SD County. For P-12 schools, testing was conducted on-site at each school, and for UCSD participants, testing was conducted on campus. Participant density was highest in regions surrounding UCSD in La Jolla, with densely tested regions extending north and south along the coast away from UCSD, and decreasing in density when moving inland and when approaching the US-Mexico border (Fig 9a). Interestingly, participants who tested positive did not necessarily follow the same density pattern as overall testing (Fig 9b). The highest density of positive participants was located around UCSD, which was expected given the large proportion of on-campus students participating in testing through the EXCITE lab. Apart from the UCSD campus, the areas with the highest proportion of positive participants included San Diego, National City, and Chula Vista, all located south of UCSD. Areas such as Encinitas and Solana Beach, to the north of UCSD, were heavily tested, but represented a disproportionately small portion of positive participants. Conversely, Chula Vista was not heavily tested, but represented a disproportionately large portion of positive participants. There are many possible explanations for this discrepancy. Median household income (2019) in Encinitas and Chula Vista was $116,022 and $81,272, respectively (https://www.census.gov/). A previous study found that a decrease of $10,000 median household income in New York City was correlated with a 1.6% increase in COVID-19 positivity rate (55). If this correlation is applied to San Diego County, we would expect positivity rates to be approximately 5.4% higher in Chula Vista as compared to Encinitas. This same study found an increase in COVID-19 positivity in more densely populated areas (55); these areas are more likely to have multi-family housing such as apartment buildings, where physical distancing is more challenging. In Encinitas, population density in 2010 was 3,164 people per square mile, while in Chula Vista, population density was 4,915/sq mi (https://www.census.gov/). Chula Vista also has significantly more Black/African American and Hispanic residents as compared to Encinitas, and both of these communities have been identified by previous studies as being at higher risk for COVID-19, as discussed above.

**Fig 9.**
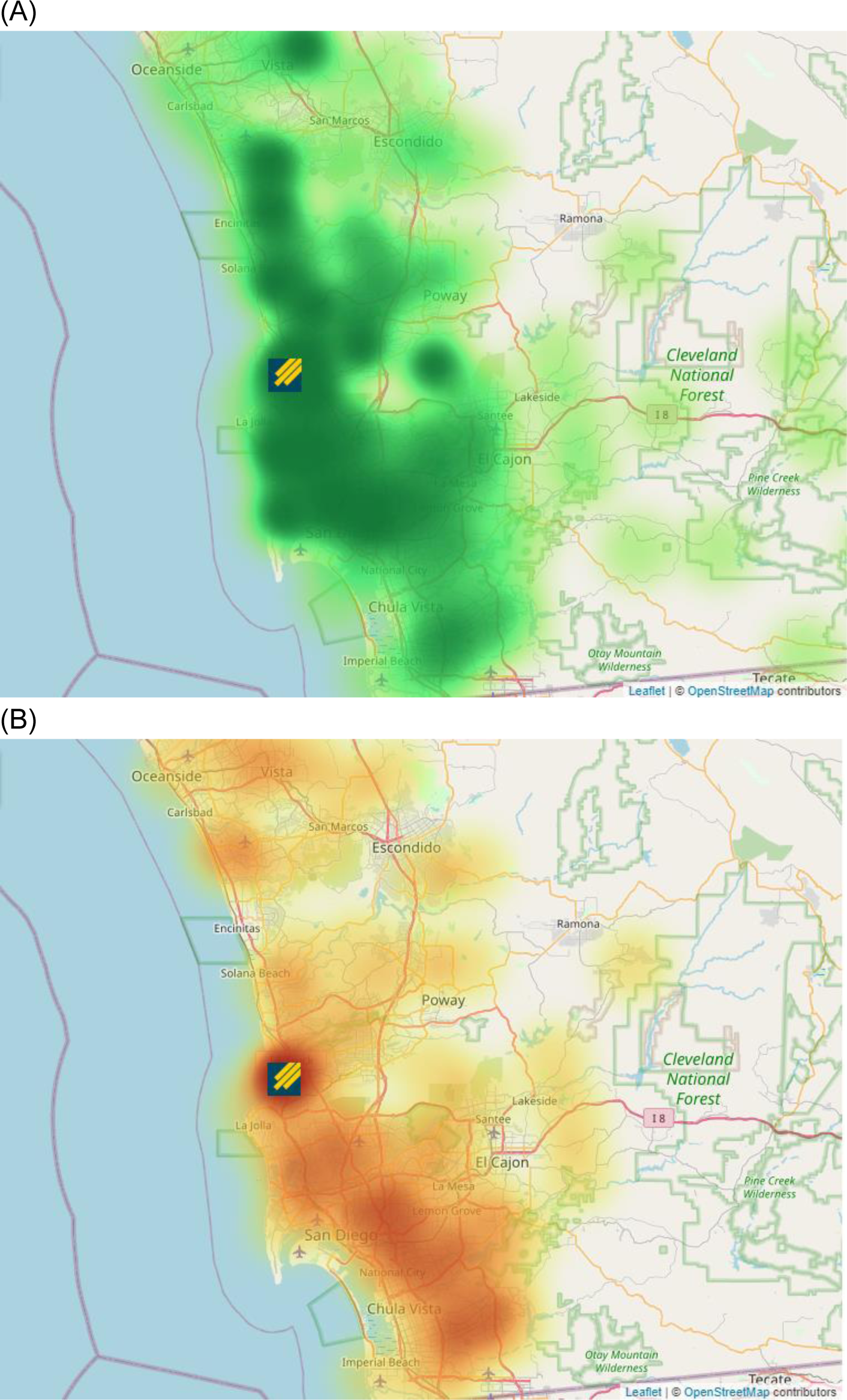
Zip codes of residence of (A) all UCSD/P-12 school participants tested through the EXCITE lab, and (B) UCSD/P-12 school participants who tested positive for COVID-19 through the EXCITE lab. The deepness of color is proportionate to the number of participants who chose that zip code as their area of residence. Zip codes from outside of San Diego County were assumed to belong to UCSD students living on-campus who provided their home address, and were re-assigned as such. The icon on both images represents the location of the UCSD campus.

#### Return to Learn at UCSD

During the fall 2020 quarter, UCSD housed 9,129 on-campus students, and COVID-19 testing on a bi-weekly basis was mandatory for any students who lived in campus owned housing or came to campus for classes. Student athletes were tested weekly when training, and more often when competing. During the fall quarter, 6% of classes were taught in-person, with a maximum in-person class size of 50, but this was reduced to 2% of classes in-person mid- quarter as SD County restrictions on indoor teaching were implemented. Following County restrictions on indoor teaching, all in-person classes were moved to outdoor settings, with large tents acting as outdoor classrooms. Testing was provided free-of-charge to students, faculty, and staff, and while testing was not mandatory for faculty and staff, it was recommended. High-volume testing sites for students moving on-campus were set up at the beginning of each quarter, and student move-in was staggered to allow for adequate testing and social distancing. Testing at term start among students moving/returning to campus-owned housing was provided at days 1 and 10 in the fall quarter. Sequestration (masking in all areas including in residences with the exception of when inside a single bedroom or bathroom) was implemented during the move in testing period.

Students living on-campus who tested positive at term start or at any point thereafter were given options on how to self-isolate for the required 14 days before being allowed to move into/return to their residential suites. Many students availed themselves of the temporary isolation housing that was provided on campus, while others chose to return home to isolate, and some, mainly graduate students, chose to isolate in their campus residence with their families or alone (this option was not presented to students living in shared housing). Students living off-campus who tested positive could remain in their homes if they could effectively isolate themselves from others, or were provided isolation housing on campus if desired.

Masking was required in all public spaces on campus, both indoors and outdoors, and beginning mid-way through the fall quarter, students were allowed to gather in groups of three for up to two hours at a time, provided everyone was outdoors, masked, and socially distanced. At certain points throughout the quarter, cases rose and students were required to sequester temporarily. Bi-weekly asymptomatic testing was accomplished via the installation of vending machines throughout campus that supplied COVID-19 testing kits containing a collection tube pre-filled with media, a swab, and instructions on how to self-collect, as well as drop-off bins, from which samples were collected multiple times per day for processing by the EXCITE lab. If at any point a student developed symptoms characteristic of COVID-19, they were tested via the CALM lab on campus and asked to self-isolate until their test results were returned.

Integration of the testing program with two smartphone applications - the UCSD student/employee app and the UCSD Health EPIC Electronic Health Record MyChart app - allowed for an easy method for linking test tube barcodes with student/employee ID numbers, and provided students and employees with their test results, whether they were negative or positive. Individuals with positive test results also received a telephone call from a clinical provider at UCSD Health, and were contacted daily during the isolation period. Most results were returned the day after sample collection. We believe this combination of quick turnaround time and the return of results for every sample increased compliance, because it provided continuous feedback to students and employees.

The number of students residing on campus decreased after the Thanksgiving 2020 weekend, because students who left for the holiday were incentivized (with partial refunds of housing costs) to remain at home until the winter quarter started in January 2021. This measure aimed to reduce an influx of new COVID-19 cases from students who gathered with other households over the Thanksgiving weekend. Students returning to campus from fall break were tested at days 1, 5, and 10 after return. Between Thanksgiving and the beginning of the winter quarter, weekly testing was encouraged but not mandatory, while bi-weekly testing remained mandatory.

When the winter quarter began in early January 2021, student move-in was again staggered to allow for all students to get tested and to prevent crowding; students were also asked to test themselves on days 1, 3, and 5 after move-in, after which time weekly testing was mandatory for those residing in campus owned housing or coming to campus. During the winter quarter, UCSD housed 8700 on-campus students who were required to test weekly, in addition to 1850 off-campus students who came to campus for classes, not including student athletes who came to campus for training and were required to test more frequently. During the winter quarter, 2% of classes were taught in-person, all of which were taught in an outdoor classroom setting. In addition to asymptomatic screening by the EXCITE lab, UCSD also offered symptomatic and exposure testing via the CALM lab. Together, the EXCITE and CALM labs made up the entirety of COVID-19 testing conducted on-campus at UCSD. Testing data from the CALM lab were not included in this study.

#### Return to Learn at Preschool-Grade 12 Schools

The EXCITE lab collaborated with 11 private schools and a group of 13 public schools in SD County to provide repeated screening of students, teachers, and staff. These schools offered different in-person learning schedules, with some offering in-person learning five days a week and others alternating between in-person and remote learning, with a portion of students attending in-person learning on different days. During the timeframe this study took place, the large majority of public schools in SD County were fully remote. However, some schools were allowed to remain open for in-person learning on a limited basis, based on the CDC’s social vulnerability index and the California Department of Public Health’s (CDPH) Healthy Places Index (HPI). Public schools that were in the lowest HPI quartile were selected, as they represented students from low socio-economic status communities, including many immigrant, refugee, and other socially vulnerable communities. Not all students from these communities participated in on-site learning; schools offered this choice to children who were struggling with remote learning due to their housing situation (homelessness, overcrowded housing, poor wifi) and/or those who were struggling academically. For public schools, testing was offered weekly but was not mandatory, and compliance was lower for students than for staff at these schools, with an average of 68% of students and 92% of staff who consented to testing. For most private schools, testing was mandatory in order to attend in-person learning, but remote learning was also available to students. The testing schedule varied for each private school, from weekly testing to exposure-based testing, but most schools required students and staff to provide a negative test before returning to school after breaks and holidays (Table S5). Schools used a variety of safety and risk mitigation measures to attempt to ensure the health of those participating in in-person learning. We were provided general information for the public schools, and more specific information for most participating private schools, detailed in Table S5 and summarized below.

At the private schools for which we obtained details of their risk mitigation strategies, cohort size ranged from 9-14 students up to the entire grade level. For at least one private school, a single positive test within a cohort meant that the entire cohort stayed at home for remote learning for two weeks, and contact tracing was used to determine any possible exposures outside the cohort. Because this method was quite disruptive for the students, administrative staff at this school recommend creating smaller cohort sizes. Desks in classrooms were placed at a distance of 4-6 ft (1.2-1.8 m), and while the established recommended distance is 6 ft (1.5 m), a recent report that has been adopted by the CDC suggests that a 3 ft (0.9 m) spacing may be just as effective at preventing the spread of SARS-CoV-2, as long as everyone is masked (56). Masking was required at all times, with exceptions for very young children and when eating (physically distanced, outside) and napping (for young children). For most schools, indoor classrooms were either retrofitted with portable home-made HEPA air filtration systems or had upgraded HVAC systems installed with higher-quality air filters. Different schools employed different indoor/outdoor instruction methods, with some schools teaching exclusively indoors, some teaching almost exclusively in outdoor classrooms, and others using a combination, emphasizing mandatory time outdoors. We note that outdoor instruction or mandatory outdoor time may not be feasible everywhere, especially in the winter months. One private school described one-way walking traffic set up throughout the campus, as well as staggering student drop-off and pick-up to prevent crowding. Different schools used different methods of contact tracing and daily symptom checking, and all six private schools that provided details of their symptom screening policies mentioned using dedicated applications, including Emocha, SchoolPass, and ProCare. These schools also implemented temperature checks upon arrival on campus. At the presence of any symptoms of illness, students and staff were required to switch to remote teaching until symptoms resolved.

At the public schools, the following extra measures were taken to ensure the health and safety of students and staff, following CDC and CDPH guidance: mandatory masking for students and staff; grouping students into cohorts to minimize interactions; 6 ft (1.5 m) distance between desks; scheduled drop-off and pick-up times; heightened ventilation in classrooms; increased sanitization and hand-washing protocols; daily symptom checks and mandatory two-week at-home quarantines for students who present symptoms of illness; restricting parents/guardians and volunteers from campus; restricting the sharing of materials between students. For most of the public schools, a single positive test within a cohort resulted in the entire cohort returning to remote learning for two weeks.

## Conclusions

In this study, we developed a high-throughput semi-automated pipeline for RT-qPCR detection of SARS-CoV-2, with scalable capacity and rapid turnaround times that was used for large-scale repeated asymptomatic screening on an individual basis. This pipeline was first used to test more than 6,000 healthcare workers and first responders in San Diego, California in the spring of 2020. The pipeline was then modified in the fall of 2020 and used to establish a dedicated CLIA-certified COVID-19 testing lab at the University of California, San Diego, allowing students and staff to return to campus safely by providing repeated asymptomatic screening. Testing was expanded to include firefighters and some preschool-grade 12 schools across San Diego County. Thus far, we have tested more than 150,000 nasal swabs from over 28,000 individuals. More than half of participants who tested positive reported no symptoms at the time of testing, highlighting the importance of asymptomatic/pre-symptomatic screening. The presence of symptoms was significantly correlated with higher viral load. Hispanic and Black/African American participants from UCSD and partnering schools were more likely to test positive, highlighting the disproportionate toll the COVID-19 pandemic has had on already-marginalized populations within the US. At UCSD and at public schools, students were more likely to test positive for COVID-19 than staff. However, in private schools, students were less likely to test positive than staff. No correlation between age/sex and viral load was observed.

We note that the results reported here were obtained during a time period (April 17, 2020 to February 5, 2021) when COVID-19 vaccination rates in San Diego were less than 10%. Our results suggest that schools ranging from preschool to university may be opened safely, even without vaccination, when proper health and safety measures are implemented, such mandatory masking, increased desk spacing, reduced cohort size, and repeat testing.

## Data Availability

Full datasets, as well as scripts for data analysis and visualization used in this manuscript, are made available at https://osf.io/3fguz/.

## Acknowledgements

We would like to acknowledge the San Diego COVID Research Enterprise Network (SCREEN) Initiative for engineering a 3D printable NP swab and scaling to meet the immediate diagnostic demands of the SEARCH study. We thank the RCHSD employees and leadership who supported the SEARCH initiative by providing their time and effort to test participants at multiple testing sites. We would like to thank the following members of the Altman Clinical & Translational Research Institute (ACTRI) for their assistance creating Figs: Melissa Generoso, Perry Shipman, Nguyen Trieu. We would like to thank the following people from the University of California San Diego (UCSD): Dr. Mana Parast from the Sanford Consortium of Regenerative Medicine for the use of her QuantStudio 5 qPCR machine; the members of the UCSD Return to Learn (RTL) program; Stosh Ozog, Joanna Coker, and Denis Smirnov from the UCSD School of Medicine. We would like to thank the following people from our partnering healthcare facilities during the SEARCH study: Cassidy Callahan, Joey Principato, and Kyle McBride from Rady Children’s Hospital, San Diego; Rowena Carino from the Department of Pathology, Scripps Clinic; Aaron Fort, Andrew Ruoff, Jessica Malley, Jamie Rullman, Amanda Beggs, and Gracie Gagliano from the Department of Surgery, Division of Transplantation, Scripps Clinic.

## Funding

Funding for the SEARCH study was provided by: the Scripps Health COVID-19 Research Fund to CM and Scripps Health collaborators (https://www.scripps.org/); a personal donation by the Shekhter Family to the SEARCH study initiative, A Centers for Disease Control and Prevention (CDC) Broad Agency Announcement (BAA) contract to KGA (contract #75D30120C09795), and departmental funds from the UCSD Department of Obstetrics, Gynecology, and Reproductive Sciences to LCL (https://medschool.ucsd.edu/som/obgyn/Pages/default.aspx). The funders had no role in study design, data collection and analysis, decision to publish, or preparation of the manuscript.

## Competing Interests

Anterior nares swab samples for clinical validation of the Expedited COVID-19 Identification Environment (EXCITE) Laboratory Developed Tests (LDT) were donated by Helix Opco, LLC. We disclose that Dr. Laurent’s spouse is an employee of Helix Opco, LLC.

## Supplemental Figs

**Fig S1.**
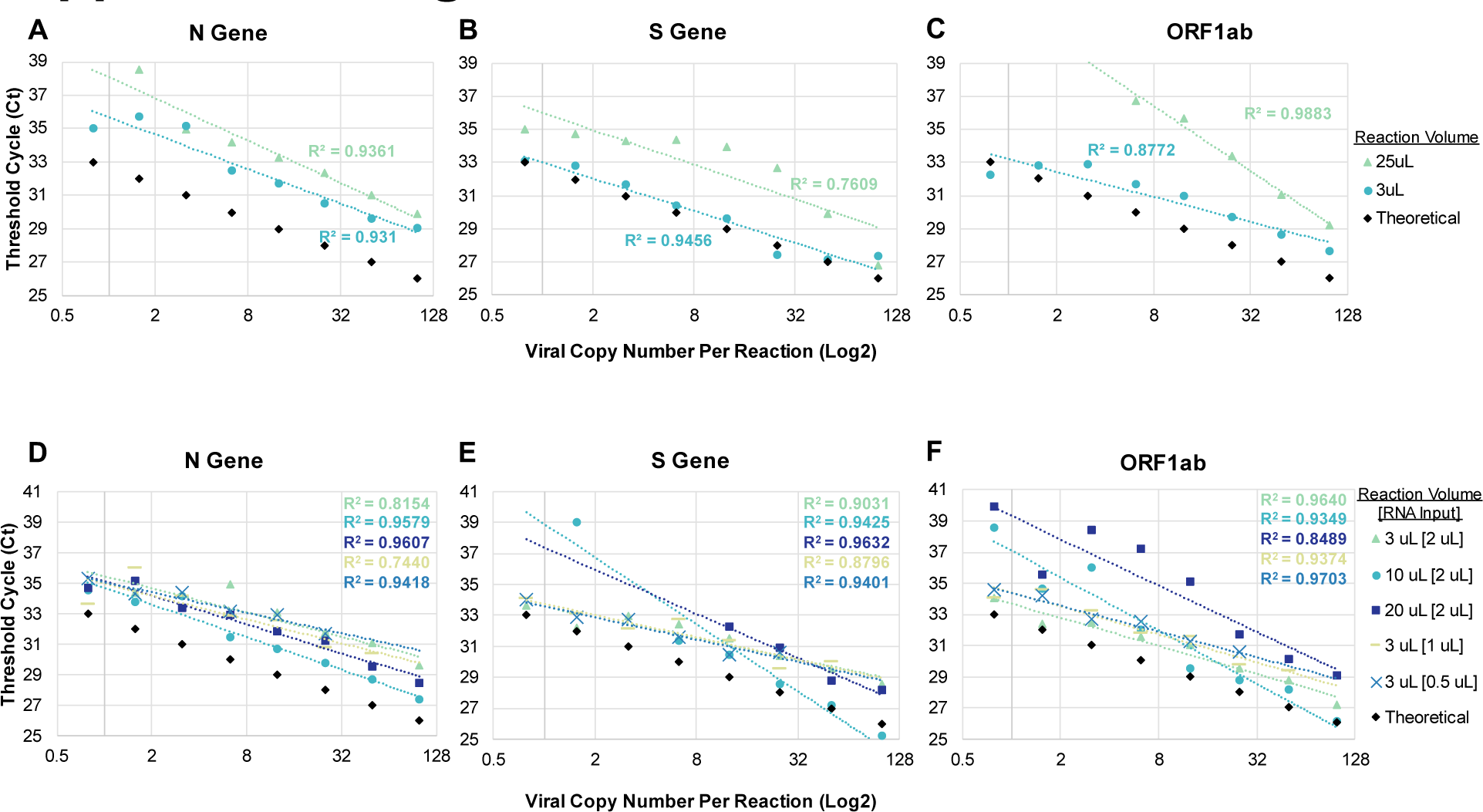
(A-C) RT-qPCR performance of miniaturized (3 µL) and full-scale (25 µL) reactions as compared to theoretical results, for three targeted regions of the SARS-CoV-2 genome: (A) N Gene; (B) S Gene; (C) ORF1ab. (D-F) RT-qPCR performance of different reactions (varied total reaction volume and RNA input volume) as compared to theoretical results, for three targeted regions of the SARS-CoV-2 genome: (A) N Gene; (B) S Gene; (C) ORF1ab. Every point in each graph represents the average of 3 technical replicates.

**Fig S2.**
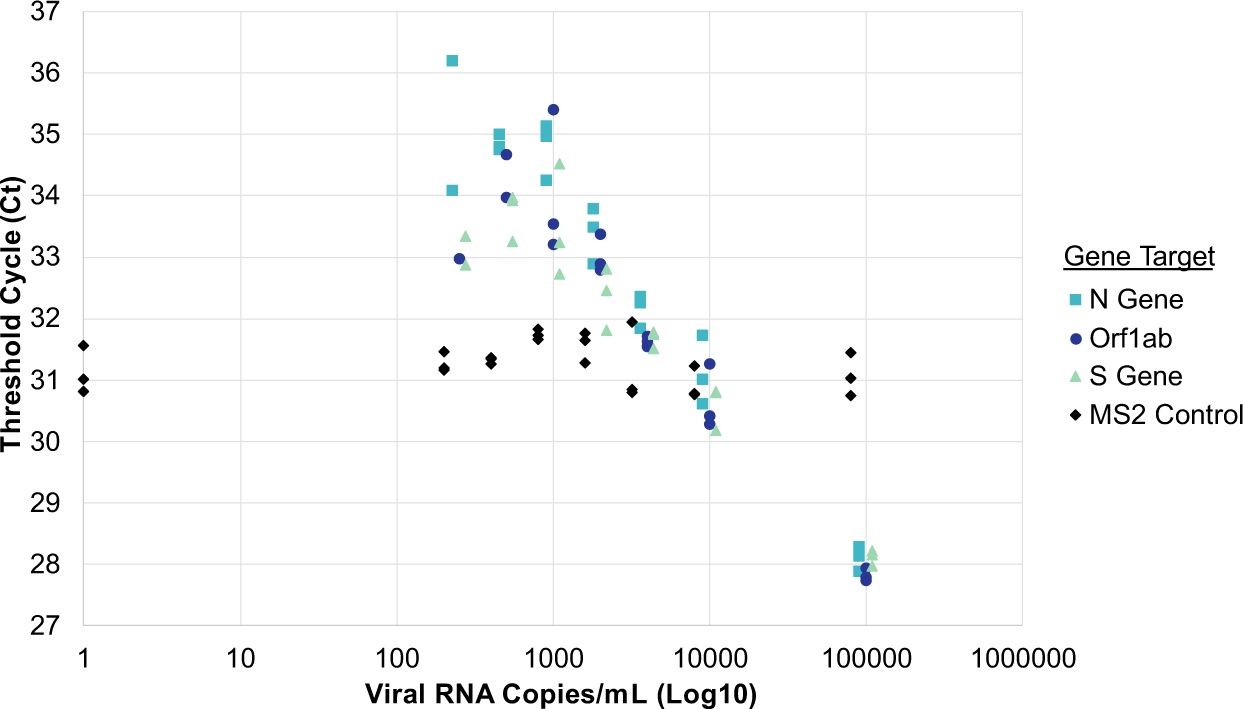
Limit of detection of SARS-CoV-2 viral RNA at different concentrations, with viral RNA spiked into negative NP control samples prior to RNA extraction. Each point represents a single replicate reaction.

**Fig S3.**
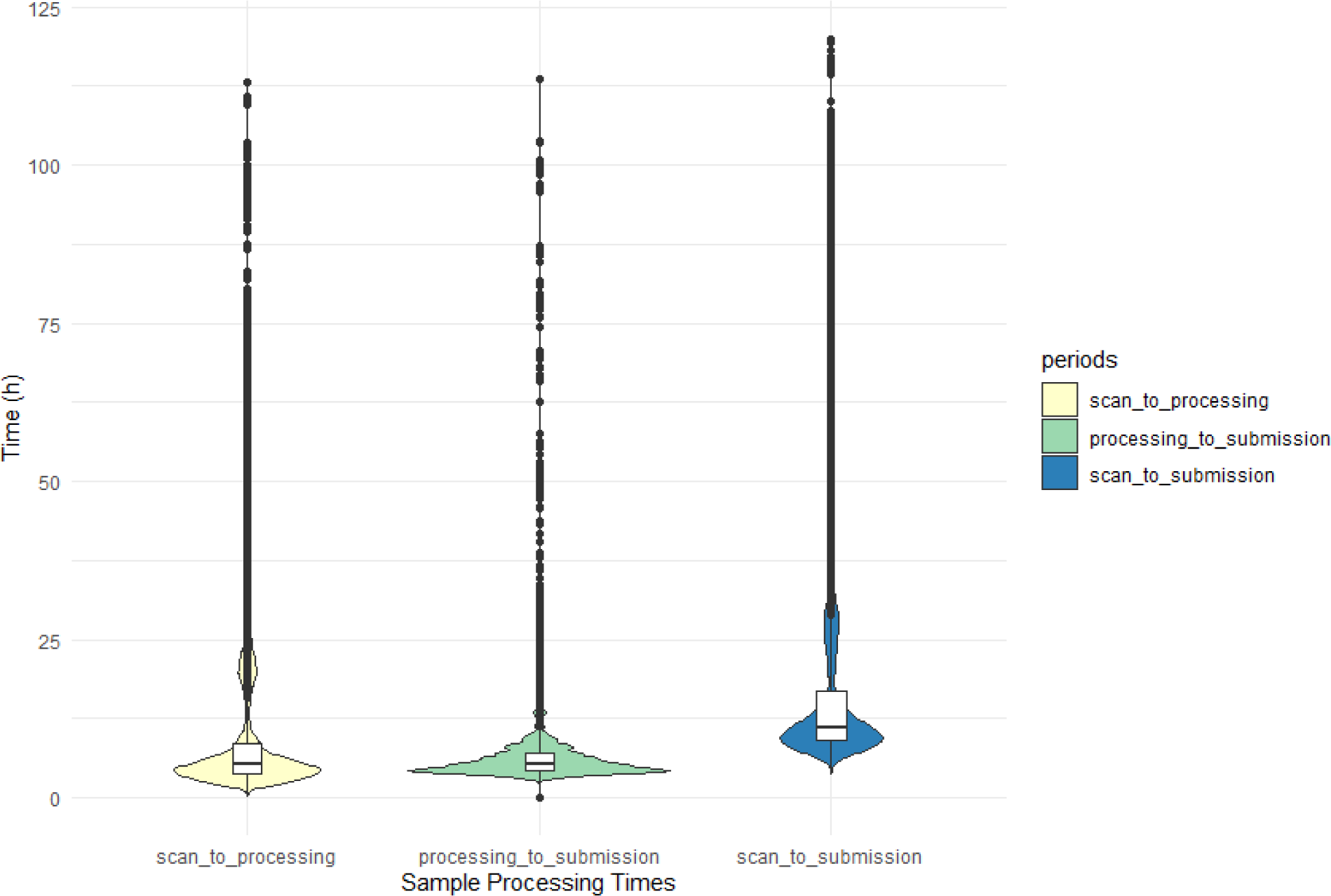
Sample processing times for samples at the EXCITE lab (n = 144,971). “Scan_to_processing” refers to the time from when a participant scans a tube barcode for self-collection to when the sample is received by the EXCITE lab and begins processing. “Processing_to_submission” refers to the time it takes to process the sample by the EXCITE lab, from the time the sample is received to the time the result is returned. “Scan_to_submission” refers to the sum of the two previous times, the total time it takes to receive results. Samples that were recorded as taking longer than five days to process were considered technical errors and were removed from this plot, along with samples with missing data for one or more of the timepoints.

**Fig S4.**
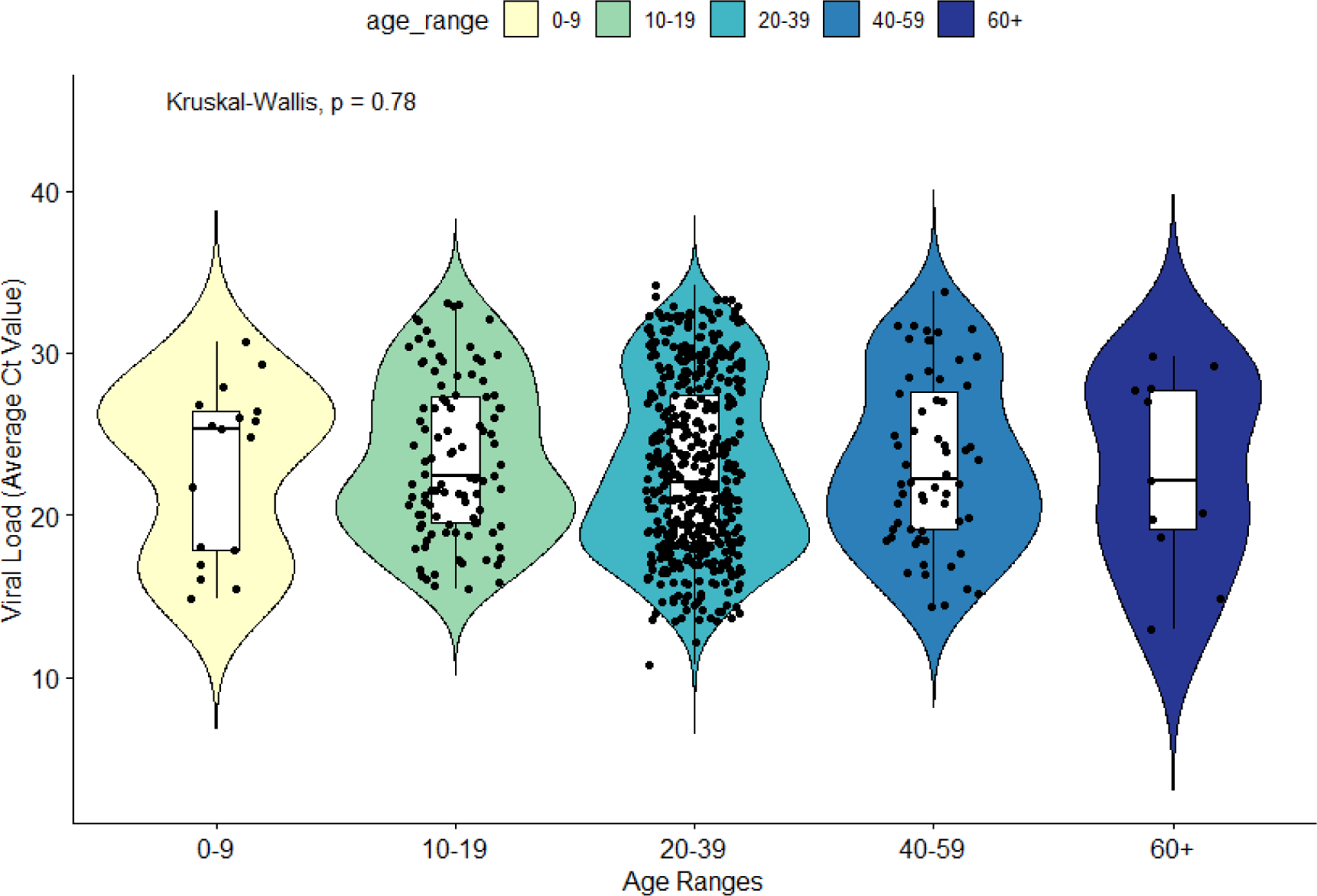
Viral load (estimated by average Ct value) of COVID-19 positive individuals, separated by age group. A Kruskal-Wallis test indicated no significant difference in viral load among age groups (*p* = 0.78).

**Fig S5.**
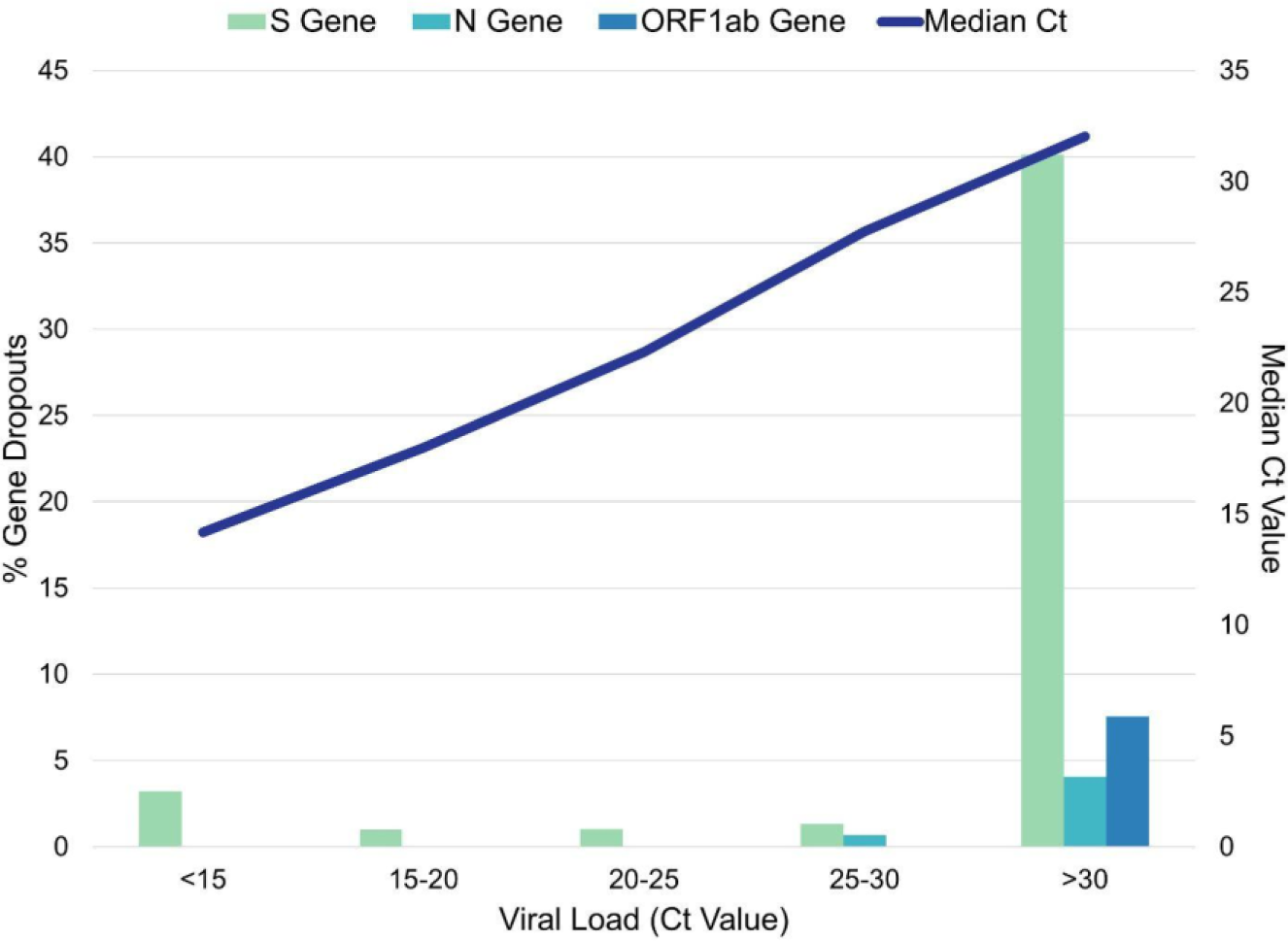
SARS-CoV-2 viral gene dropouts during RT-qPCR detection as a function of median viral load (Ct value).

**Fig S6.**
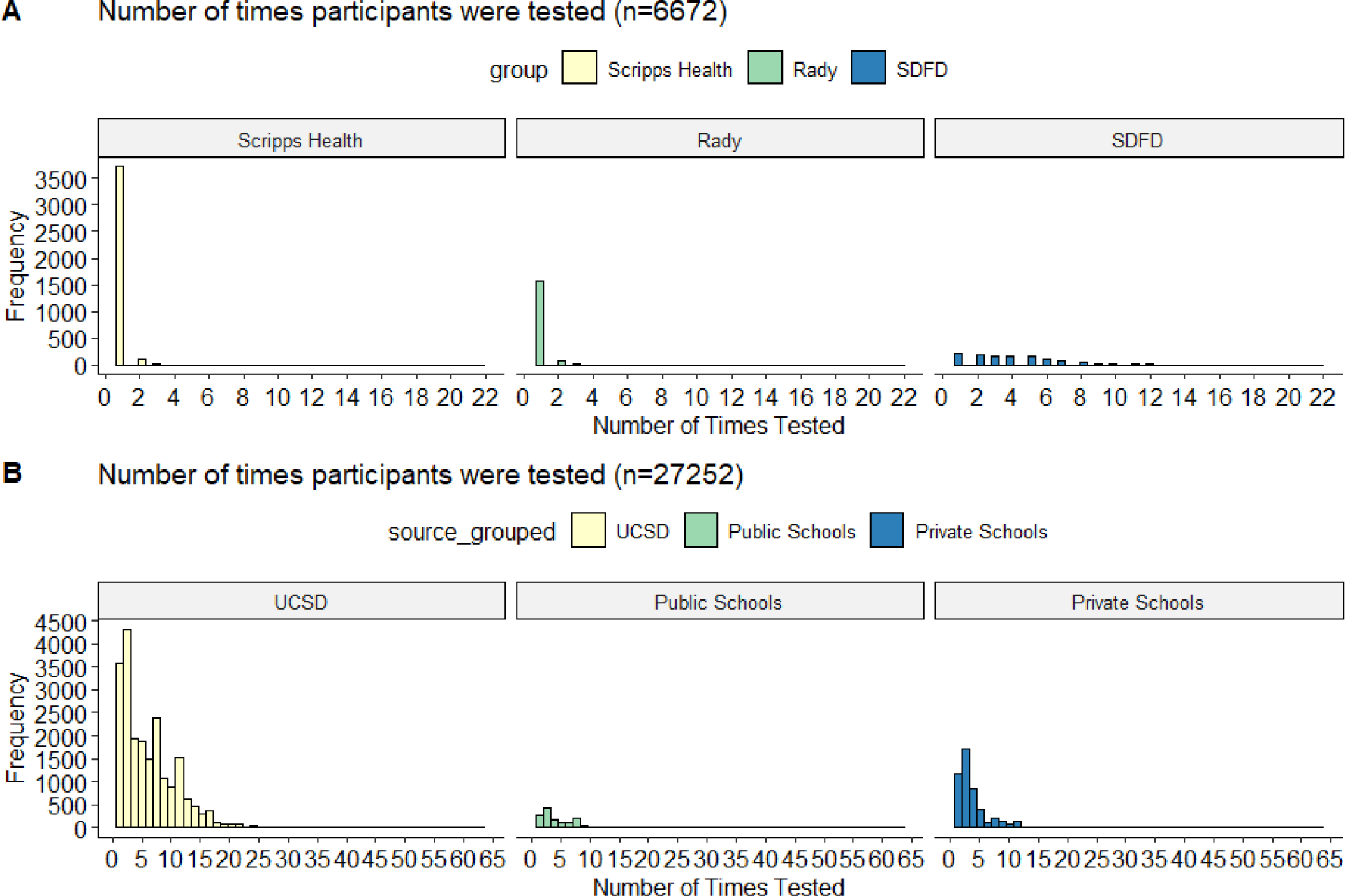
The number of times (A) SEARCH study participants and (B) EXCITE lab participants chose to be tested for SARS-CoV-2. Healthcare workers from Scripps Health and Rady systems were tested between April 17 and June 30, 2020 via the SEARCH study. Students, faculty, and staff from the University of California, San Diego (UCSD) and from public and private schools in San Diego County were tested between Sept 15, 2020 and February 5, 2021 via the EXCITE lab. San Diego Fire-Rescue Department (SDFD) participants were tested via both the SEARCH study and the EXCITE lab during the same timeframes.

## Supplemental Tables

**Table S1.** Comparison of 3D-printed and commercially-sourced nasopharyngeal swabs. In total, 25 pediatric patients and nine adult patients were swabbed to perform the validation of the 3D- printed swabs. Of the 25 pediatric samples, 22 were tested using a viral respiratory panel, and three were tested using a COVID-19 test. All nine adult samples were COVID-19 positive; however, only four had both 3D-printed and commercial swabs.

|  | Commercial Swab | 3D-Printed Swab |
| --- | --- | --- |
| **Pediatric** |  |  |
| Respiratory Viral Panel |  |  |
| Negative | 16/16 (100%) | 16/16 (100%) |
| Positive (single) | 3/5 (60%) | 5/5 (100%) |
| Positive (multiple) | 1/1 (100%) | 1/1 (100%) |
| COVID-19 negative | 2/2 (100%) | 2/2 (100%) |
| COVID-19 positive | 1/1 (100%) | 1/1 (100%) |
| **Adult** |  |  |
| COVID-19 positive (in parallel) | 2/4 (50%) | 4/4 (100%) |
| COVID-19 positive (3D swab only) | Not available | 5/5 (100%) |

**Table S2.** Technical validation of anterior nares swabs in different media for SARS-CoV-2 detection: PrimeStore® Molecular Transport Medium (MTM media/ANM sample type); Mawi DNA Technologies iSWAB Microbiome buffer (Mawi media/ANW sample type), and 5% sodium dodecyl sulfate (SDS media/AND sample type). Limit of detection for triplicate contrived samples containing different concentrations of viral particles (VP) per mL (250 - 32,000). Rows in bold indicate the limit of detection for that media type. Three SARS-CoV-2 viral genes were targeted for amplification (N gene, Orf1ab, and S gene), as well as an internal control (MS2). A minimum of 2/3 viral gene targets were required to amplify within each replicate to be considered a positive result.

|  |  | N Gene |  |  | Orf1ab |  |  | S Gene |  |  | MS2 |  |  |
| --- | --- | --- | --- | --- | --- | --- | --- | --- | --- | --- | --- | --- | --- |
|  | VP/mL | 1 | 2 | 3 | 1 | 2 | 3 | 1 | 2 | 3 | 1 | 2 | 3 |
| ANM | 32000 | 28.56724 | 27.30496 | 29.05796 | 28.123 | 26.74671 | 28.40087 | 28.23621 | 26.90927 | 28.74655 | 30.88804 | 30.39208 | 31.40189 |
|  | 16000 | 28.59086 | 29.10027 | 30.37453 | 27.87927 | 28.2653 | 29.04837 | 28.28125 | 28.63926 | 30.22964 | 30.47556 | 30.3301 | 32.33632 |
|  | 8000 | 29.622 | 29.6161 | 31.03946 | 28.8252 | 28.9988 | 30.13729 | 29.59232 | 29.40543 | 30.42216 | 30.80075 | 30.81684 | 32.18524 |
|  | 4000 | 30.55292 | 29.34379 | 31.57382 | 29.06067 | 28.92757 | 30.08623 | 30.36565 | 30.0086 | 30.64664 | 30.94963 | 31.10299 | 32.24866 |
|  | 2000 | 29.79179 | 30.78633 | 32.3596 | 29.65664 | 29.51823 | 30.53481 | 30.80253 | 30.60761 | 32.05163 | 30.91378 | 30.87834 | 32.21166 |
|  | 1000 | 31.95534 | 31.63872 | 32.23191 | 30.48554 | 30.74818 | 31.60106 | 31.6986 | 30.66024 | 31.82981 | 31.09695 | 30.54766 | 31.51518 |
|  | **500** | **31.8007** | **32.17833** | **32.93955** | **31.25087** | **30.67037** |  | **31.43116** | **31.69549** | **32.06826** | **31.05981** | **31.16852** | **31.70178** |
|  | 250 | 32.91542 | 32.62193 |  | 31.56975 | 31.78306 | 32.0334 | 32.01559 | 31.93159 |  | 30.60415 | 30.89187 | 32.10643 |
| ANW | 32000 | 27.6949 | 27.62062 | 28.6829 | 26.75763 | 27.07014 | 27.86881 | 27.33813 | 27.60611 | 28.84602 | 29.7765 | 29.94439 | 30.21699 |
|  | 16000 | 27.77016 | 27.98963 | 30.38937 | 27.51569 | 27.56042 | 28.88784 | 28.21654 | 28.12109 | 29.61706 | 30.10374 | 30.00491 | 30.40088 |
|  | 8000 | 29.58683 | 28.77404 | 29.10565 | 28.46152 | 28.83664 | 28.64296 | 29.12567 | 29.02891 | 29.54257 | 30.49041 | 30.77581 | 30.56796 |
|  | 4000 | 30.67838 | 30.55936 | 30.49048 | 28.79253 | 29.14261 | 29.4781 | 30.15295 | 29.68094 | 29.89257 | 30.60305 | 30.637 | 30.54589 |
|  | 2000 | 30.89805 | 31.18324 | 32.13199 | 30.06639 | 29.95144 | 30.58552 | 31.16762 | 30.60806 | 31.78862 | 30.70745 | 30.53986 | 31.03023 |
|  | 1000 | 30.69853 | 31.70212 | 33.70245 | 30.83089 | 31.16146 | 31.39502 | 31.41211 | 31.37258 | 32.74824 | 30.87244 | 30.66293 | 31.0696 |
|  | 500 | 33.08861 | 32.74819 | 33.7467 | 31.24067 | 31.50742 | 32.27259 | 32.32956 | 31.97728 | 32.70442 | 30.81637 | 30.69168 | 31.30658 |
|  | **250** | **32.69856** |  | **33.85752** | **31.97604** | **31.24246** | **32.41334** | **31.77786** | **31.17495** |  | **30.5527** | **30.24898** | **30.31604** |
| AND | 32000 | 26.56684 | 26.48251 | 26.43064 | 26.01545 | 25.92576 | 25.85998 | 26.34711 | 26.31556 | 26.20625 | 30.40321 | 30.30017 | 30.44547 |
|  | 16000 | 27.63647 | 27.65226 | 27.66501 | 26.83035 | 26.90872 | 27.02284 | 27.21644 | 27.42337 | 27.63582 | 30.42951 | 30.16785 | 30.07215 |
|  | 8000 | 28.48938 | 28.46131 | 28.30104 | 27.79799 | 27.74197 | 27.61454 | 27.76735 | 28.23767 | 27.80854 | 30.27699 | 30.45726 | 30.03353 |
|  | 4000 | 28.95999 | 28.80925 | 28.81392 | 28.00529 | 28.47926 | 28.04047 | 28.37351 | 28.91348 | 28.77715 | 30.12698 | 30.34609 | 29.99662 |
|  | 2000 | 30.77094 | 30.59611 | 29.90652 | 29.35722 | 29.33499 | 29.5718 | 29.733 | 29.9439 | 29.90099 | 30.43141 | 30.74416 | 30.84527 |
|  | 1000 | 30.85673 | 30.78722 | 31.12153 | 29.11394 | 29.77203 | 29.98141 | 29.87768 | 29.8279 | 30.48528 | 30.46241 | 30.82404 | 30.39688 |
|  | 500 | 31.41748 | 32.30139 | 31.82398 | 29.82488 | 30.51586 | 30.76725 | 30.77686 | 30.89876 | 30.61647 | 30.47791 | 30.87041 | 30.89983 |
|  | **250** | **32.77331** | **33.0288** | **32.94244** | **29.98244** | **30.56765** | **30.93435** | **31.60111** | **31.51474** | **31.75622** | **30.88731** | **30.77039** | **30.52082** |

**Table S3.** Technical validation of anterior nares swabs in different media for SARS-CoV-2 detection: PrimeStore® Molecular Transport Medium (MTM media/ANM sample type); Mawi DNA Technologies iSWAB Microbiome buffer (Mawi media/ANW sample type), and 5% sodium dodecyl sulfate (SDS media/AND sample type). A minimum of 20 contrived samples all containing the same concentration of viral particles (1000 VP/mL) were tested for each media type. Three SARS-CoV-2 viral genes were targeted for amplification (N gene, Orf1ab, and S gene), as well as an internal control (MS2). A minimum of 2/3 viral gene targets were required to amplify within each replicate to be considered a positive result, and a minimum of 19/20 samples were required to return a positive result for this technical validation step.

|  | ANM |  |  |  | ANW |  |  |  | AND |  |  |  |
| --- | --- | --- | --- | --- | --- | --- | --- | --- | --- | --- | --- | --- |
|  | N Gene | Orf1ab | S Gene | MS2 | N Gene | Orf1ab | S Gene | MS2 | N Gene | Orf1ab | S Gene | MS2 |
| Replicate 1 | 33.20503 | 31.2742 | 33.32748 | 32.13901 | 32.328 | 29.96588 | 31.68405 | 30.2813 | 30.36313 | 28.84117 | 29.74154 | 29.97011 |
| Replicate 2 | 33.11781 | 31.22991 | 31.90709 | 31.49288 | 32.665 | 30.83412 | 32.83377 | 30.47811 | 30.81601 | 30.43513 | 30.74937 | 30.34116 |
| Replicate 3 | 32.15603 | 30.20218 | 31.47673 | 31.44101 | 33.419 | 30.95764 | 31.92202 | 30.8268 | 31.02649 | 30.16447 | 30.39575 | 30.27942 |
| Replicate 4 | 32.12903 | 30.85151 | 31.2922 | 31.45474 | 33.009 | 30.14325 | 31.71588 | 30.55374 | 31.50233 | 30.96156 | 30.56672 | 30.63448 |
| Replicate 5 | 32.66456 | 30.69429 | 31.49954 | 31.66452 | 33.065 | 30.48087 | 32.26574 | 30.59831 | 30.90308 | 29.73466 | 29.95023 | 29.85332 |
| Replicate 6 |  | 30.76613 | 31.83545 | 31.09046 |  |  | 32.32624 | 30.43625 | 30.81576 | 29.68443 | 29.94711 | 29.85306 |
| Replicate 7 | 31.54116 | 30.99806 | 30.75484 | 31.44883 | 31.949 | 30.05961 | 31.15406 | 30.01067 | 31.22313 | 28.91922 | 29.96547 | 30.3383 |
| Replicate 8 | 31.26402 | 30.47032 | 30.82045 | 31.08399 | 30.934 | 30.81955 | 30.36134 | 29.98146 | 31.09156 | 29.31754 | 30.72157 | 30.03009 |
| Replicate 9 | 31.64176 | 30.93807 | 30.8849 | 30.83263 | 31.736 | 31.03576 | 31.92403 | 30.77339 | 31.06431 | 30.29006 | 30.41665 | 30.08424 |
| Replicate 10 | 32.00396 | 31.02591 | 31.52836 | 31.55012 | 33.078 | 30.57624 | 31.86759 | 30.87953 | 30.85805 | 30.70002 | 30.23393 | 30.21832 |
| Replicate 11 | 32.84344 | 30.4075 | 31.35707 | 31.26224 | 33.776 | 30.80046 | 31.88817 | 30.70583 | 30.83426 | 30.12243 | 30.41913 | 30.12273 |
| Replicate 12 | 31.63306 | 30.15641 | 31.23138 | 31.56804 | 32.900 | 30.88295 | 31.94054 | 31.03309 | 31.44029 | 30.38277 | 30.81528 | 30.20662 |
| Replicate 13 | 31.4435 | 30.99032 | 30.90512 | 31.36599 | 32.569 | 30.44113 | 32.60187 | 30.92437 | 30.65612 | 30.03494 | 29.91626 | 29.9131 |
| Replicate 14 | 31.06826 | 30.82614 | 30.56917 | 30.77931 | 33.554 | 31.75565 | 32.11547 | 30.57389 | 30.29833 | 29.73377 | 29.77454 | 29.81347 |
| Replicate 15 | 31.12846 | 30.33739 | 30.75388 | 30.90927 | 31.823 | 30.77881 | 30.93723 | 30.43053 | 31.29008 | 28.71616 | 30.271 | 29.92114 |
| Replicate 16 | 32.10682 | 30.81051 | 31.40603 | 30.78367 | 33.312 | 30.96995 | 31.25103 | 30.42333 | 30.91602 | 29.76852 | 29.95509 | 31.00376 |
| Replicate 17 | 31.30213 | 30.68539 | 31.46759 | 30.47551 | 32.177 | 30.40417 | 32.03503 | 31.32467 | 30.81853 | 30.25693 | 30.90988 | 30.38307 |
| Replicate 18 | 31.28477 | 30.36862 | 30.79565 | 30.63984 | 32.393 | 30.88316 | 31.48466 | 31.36795 | 30.81384 | 29.5703 | 30.3437 | 30.28908 |
| Replicate 19 | 31.48638 | 30.27696 | 30.66186 | 30.81963 | 32.470 | 30.78672 | 31.53987 | 31.22123 | 30.9675 | 30.30584 | 30.77847 | 29.92241 |
| Replicate 20 | 30.9396 | 30.73603 |  | 30.78493 | 32.968 | 30.75034 | 31.76518 | 31.39022 | 30.77984 | 29.19349 | 30.61045 | 30.16572 |
| Replicate 21 | N/A | N/A | N/A | N/A | N/A | N/A | N/A | N/A | 31.18732 | 30.29526 | 30.71653 | 30.28227 |
| Replicate 22 | N/A | N/A | N/A | N/A | N/A | N/A | N/A | N/A | 31.24267 | 30.011 | 30.23237 | 30.0241 |

**Table S4.** Clinical validation of anterior nares swabs in different media for SARS-CoV-2 detection: PrimeStore® Molecular Transport Medium (MTM media/ANM sample type); Mawi DNA Technologies iSWAB Microbiome buffer (Mawi media/ANW sample type), and 5% sodium dodecyl sulfate (SDS media/AND sample type). A minimum of 30 positive and 30 negative clinical samples were tested for each media type. Clinical validation involved testing each sample twice, once using the experimental test to be validated and once with an FDA Emergency Use Authorization (EUA) validated comparator. For ANM validation, the comparator was an EUA-authorized test developed by Helix LLC that also used MTM media; for ANW and AND validation, the newly-validated ANM test was used as the comparator. Three SARS-CoV-2 viral genes were targeted for amplification (N gene, Orf1ab, and S gene), as well as an internal control (MS2). A minimum of 2/3 viral gene targets were required to amplify within each replicate to be considered a positive result, and a minimum 90% sensitivity/specificity (positive/negative agreement between samples tested with the experimental and comparator tests, respectively) was required for clinical validation. Numbers in brackets represent 95% confidence intervals.

|  | ANM |  | ANW |  | AND |  |
| --- | --- | --- | --- | --- | --- | --- |
|  | Positive | Negative | Positive | Negative | Positive | Negative |
| **Comparator Positive** | 30 | 0 | 32 | 0 | 35 | 2 |
| **Comparator Negative** | 0 | 32 | 0 | 54 | 0 | 54 |
| **Sensitivity** | 100% (88.43 - 100) |  | 100% (89.11 - 100) |  | 94.59% (81.81 - 99.34) |  |
| **Specificity** | 100% (89.11 - 100) |  | 100% (93.40 - 100) |  | 100% (93.40 - 100) |  |
| **Accuracy** | 100% (94.22 - 100) |  | 100% (95.80 - 100) |  | 97.80% (92.29 - 99.73) |  |

**Table S5.**
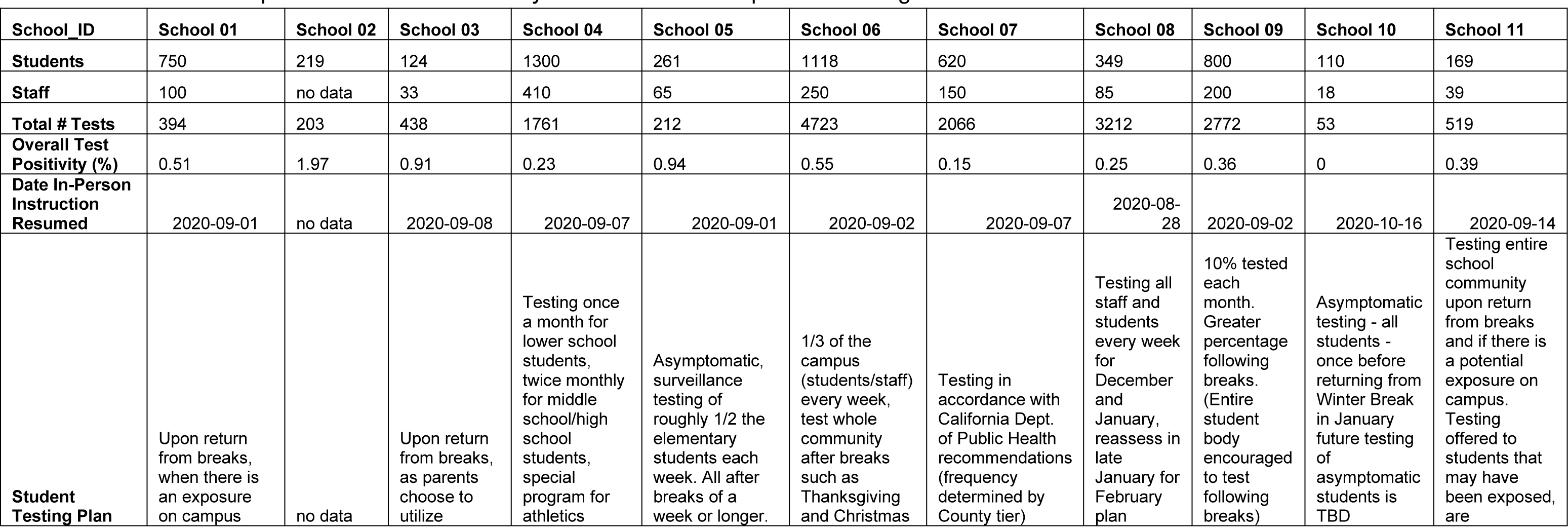
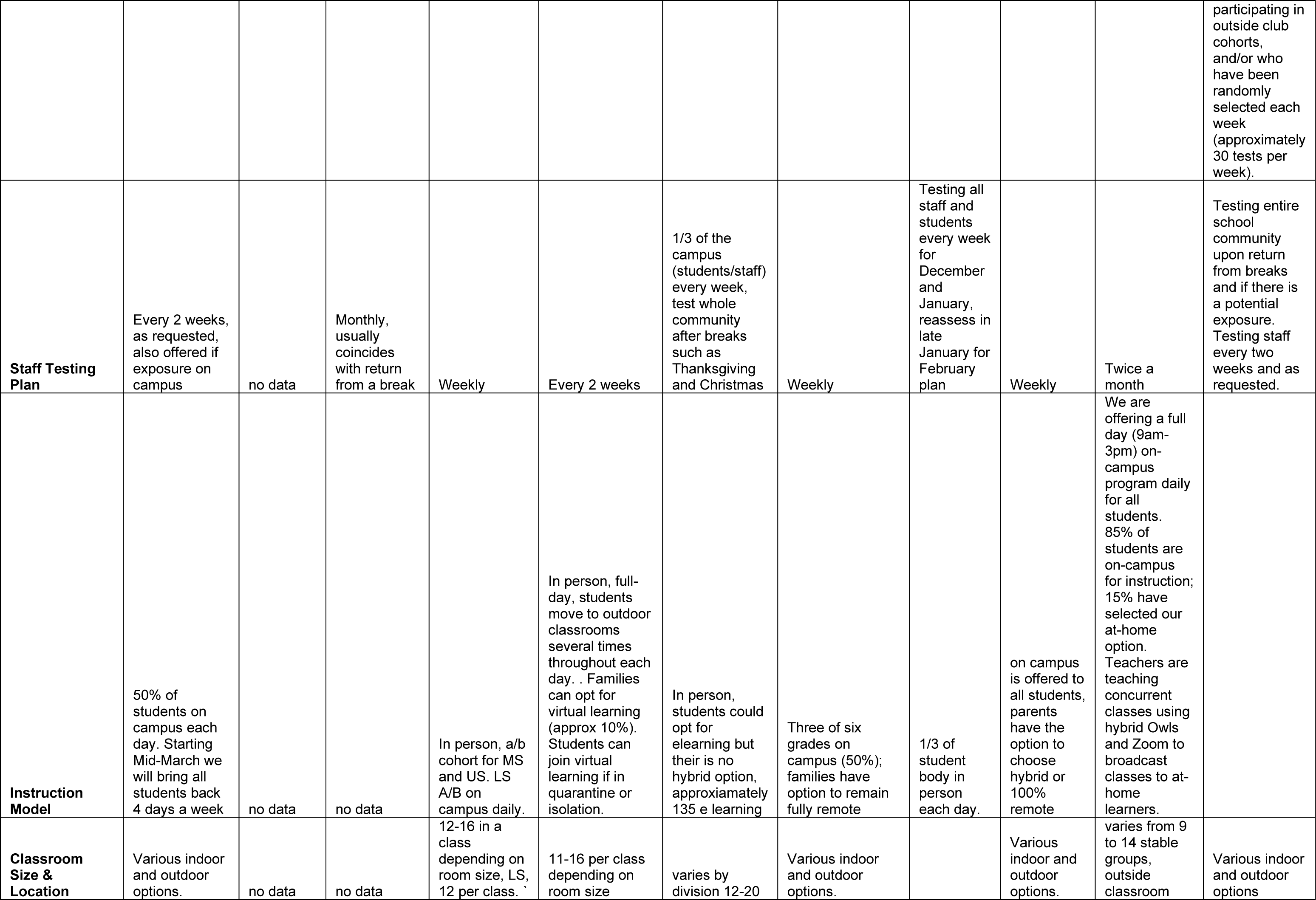
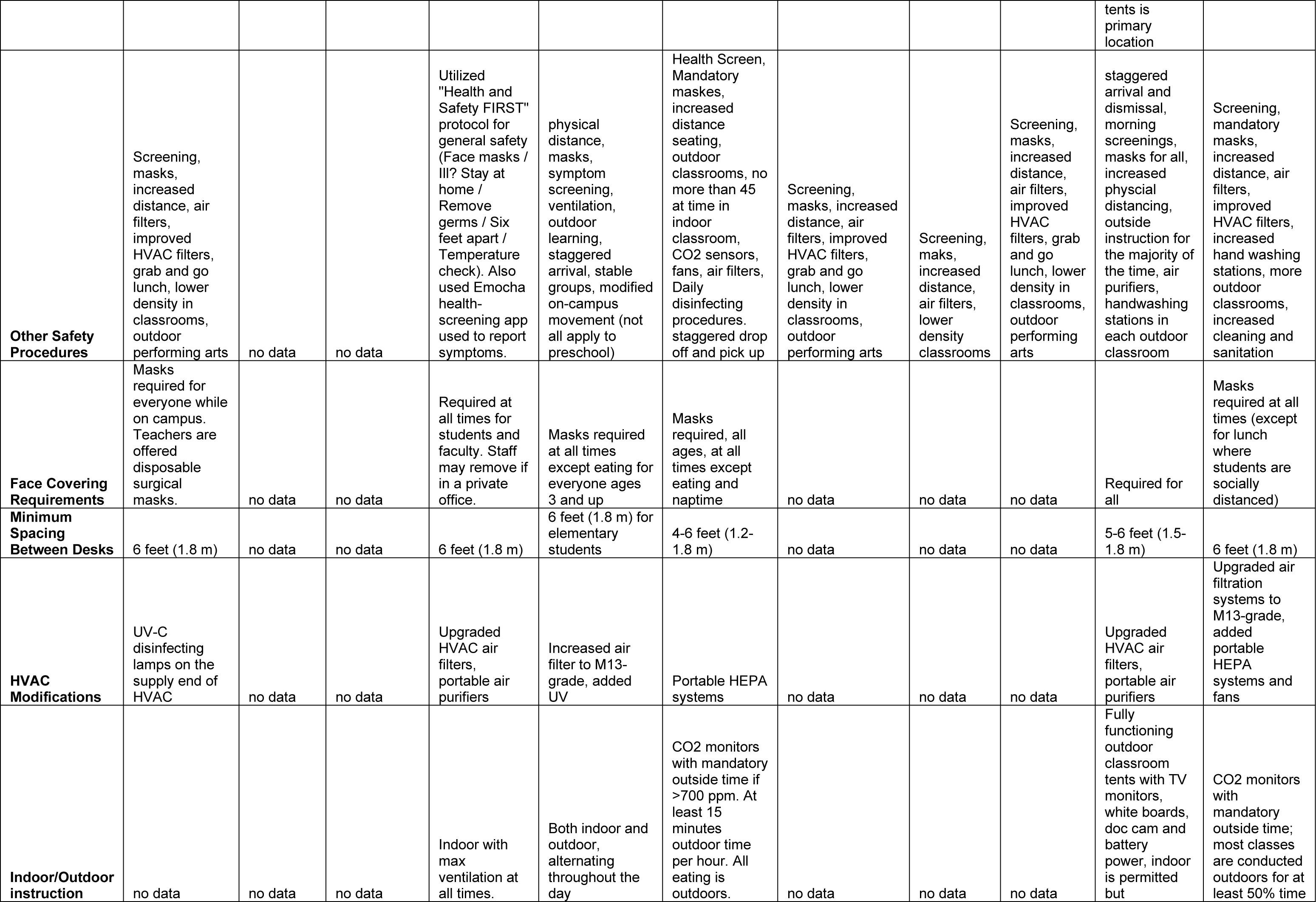
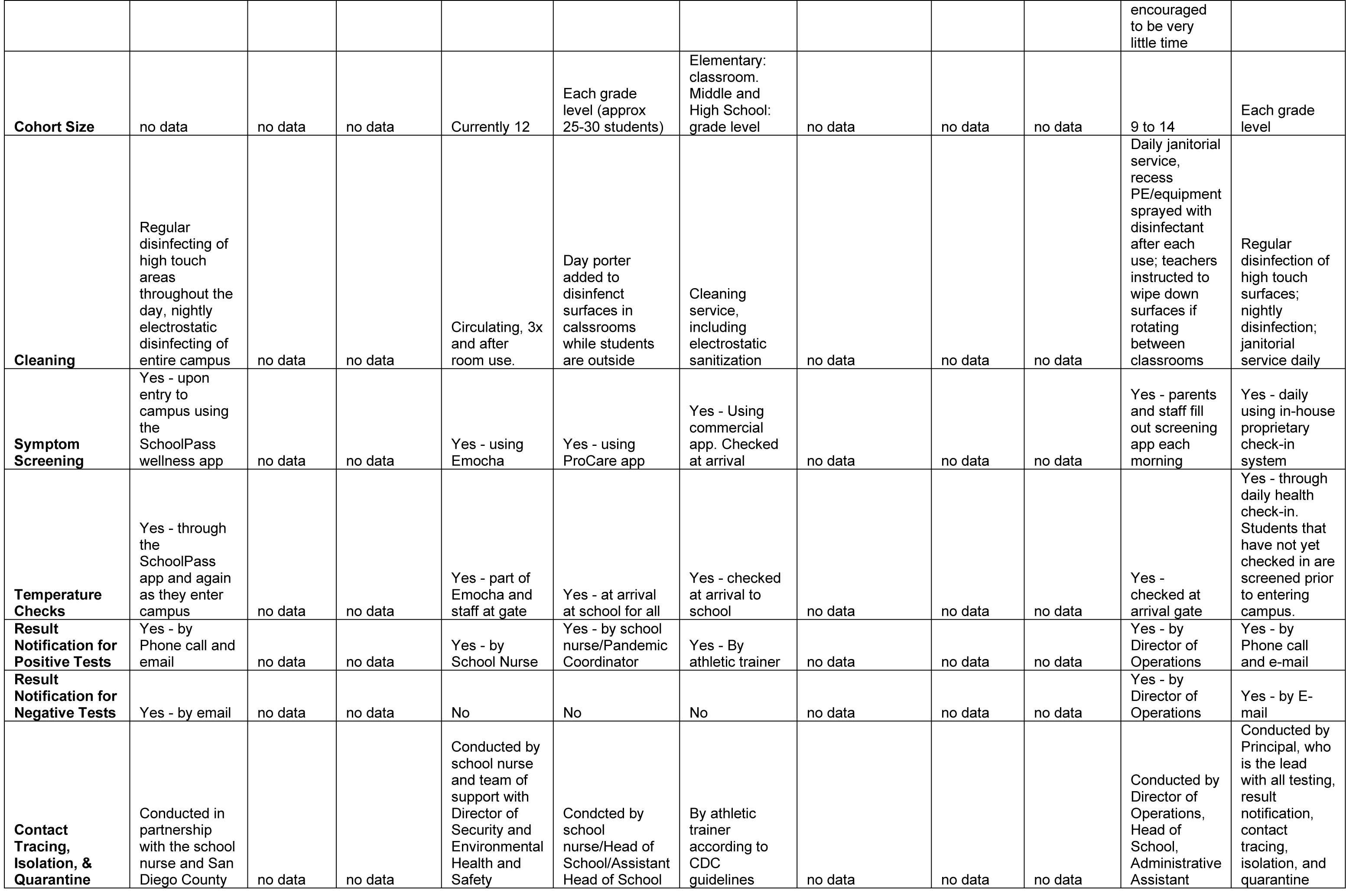
Private school partners - health and safety modifications for in-person learning.

## Notes

### Clinical Protocols

https://www.protocols.io/view/3d-printed-nasopharyngeal-swabs-with-wrapped-rayon-bemxjc7n

https://www.protocols.io/view/inspect-sample-tracking-system-bis8kehw

https://www.protocols.io/view/semi-automated-extraction-of-viral-rna-using-the-o-brqhm5t6

https://www.protocols.io/view/semi-automated-extraction-of-viral-rna-using-the-m-brcvm2w6

https://www.protocols.io/view/semi-automated-and-miniaturized-sars-cov-2-detecti-brhvm366

### Author Declarations

The University of California San Diego Institutional Review Boards (IRB) provided human subject protection oversight of the SEARCH study (IRB approval #200470). The UCSD Institutional Review Boards (IRB) provided human subject protection oversight of the data obtained by the EXCITE lab (IRB approval #210817X).

### Summary of Updates

Updated author list

